# Converging molecular evolution in acute myeloid leukaemia

**DOI:** 10.1101/2020.11.03.20222885

**Authors:** Caroline Engen, Monica Hellesøy, Tara Helén Dowling, Samuli Eldfors, Brent Ferrell, Stein-Erik Gullaksen, Mihaela Popa, Atle Brendehaug, Riikka Karjalainen, Eline Mejlænder-Andersen, Muntasir Mamun Majumder, Kimmo Porkka, Randi Hovland, Øystein Bruserud, Jonathan Irish, Caroline Heckman, Emmet McCormack, Bjørn T. Gjertsen

## Abstract

Acute myeloid leukaemia (AML) is a highly heterogeneous disease. Here, we decipher the disease composition of a single AML patient through longitudinal sampling scrutinized by high-resolution genetic and phenotypic approaches, including sequencing, immunophenotyping, *ex vivo* drug sensitivity testing and establishment of patient-derived xenograft models. Throughout the disease course we identified patterns of both divergent and convergent molecular evolution within the leukemic compartment. We identified at least six discrete leukaemia initiating cell populations, of which five were characterised by known recurrent mutations in AML. These populations partly correlated with immunophenotypically defined cell subsets, drug sensitivity profiles and population-specific potential for engraftment in immunodeficient mice. Our results indicate that the genetic and phenotypic development are closely intertwined, and that diversity in the leukaemic gene-environment likely influences disease trajectories.

**SIGNIFICANCE:** Novel therapeutic approaches in AML are characterised by targeting molecular mechanisms thought to drive leukemogenesis, but emergent evidence suggests that intra-leukemic composition may be more diverse than previously appreciated. Through in-depth genetic and phenotypic characterization of the disease course of a single AML patient, we demonstrate a high degree of inter-individual complexity that exceeds the prevailing disease conception. The temporal molecular landscape of this patient suggests that leukemogenic transitions may not be categorically monoclonal. Patterns of converging molecular evolution further imply that higher levels of biological organisation than the molecular machinery of single cells may influence leukemogenic trajectories. Disease dynamics, relational properties and causal contribution from several levels of biological organization comes into conflict with the linear monocausal explanatory model on which precision oncology is largely built. This may have implications for current precision oncology oriented prectices, including molecular categorization, molecular therapeutic targeting and predictive models.

## INTRODUCTION

Acute myeloid leukaemia (AML) is a haematopoietic stem and progenitor cell disorder, characterised by dysfunctional haematopoietic maturation and expansion of immature haematopoietic cells^1,2^. The phenotypic and genotypic variability of the disease amongst individuals is extensive, and numerous genetic^3,4^, epigenetic^5^ and transcriptional features^6^ have been associated with treatment response and disease outcome. The objective of improving outcome in AML has led to a considerable effort in stratifying patients into molecularly defined subgroups, suitable for targeted therapeutic approaches^7^. However, long-term survival rates in AML remain poor as the translation of precision haemato-oncology to durable, clinically beneficial therapies have been unexpectedly challenging^8^. Several novel compounds have been approved in the recent years, yet the clinical benefit they provide is still mostly restricted to transient partial responses^9^.

Recent technological advances have allowed increased resolution of the architecture of AML, revealing unexpected complexity. AML is now known to be composed of functionally and genetically diverse populations of cells. The relationship between these populations is dynamic and frequently fluctuate through disease courses^10-14^. This in part accounts for the current disappointments of precision oncology in AML: the targets are frequently plural, and leukemic cell composition is dynamic and evolving.

Individual reports of AML disease courses comprising detailed accounts of intra-leukemic composition and kinetics are poorly represented in the literature. In this study, we show a complex and highly dynamic process of clonal evolution in one AML patient (Figure 1A). We further synthesise a line of epistemic and translational considerations concerning the current conceptualisation and management of AML.

**Figure 1.**
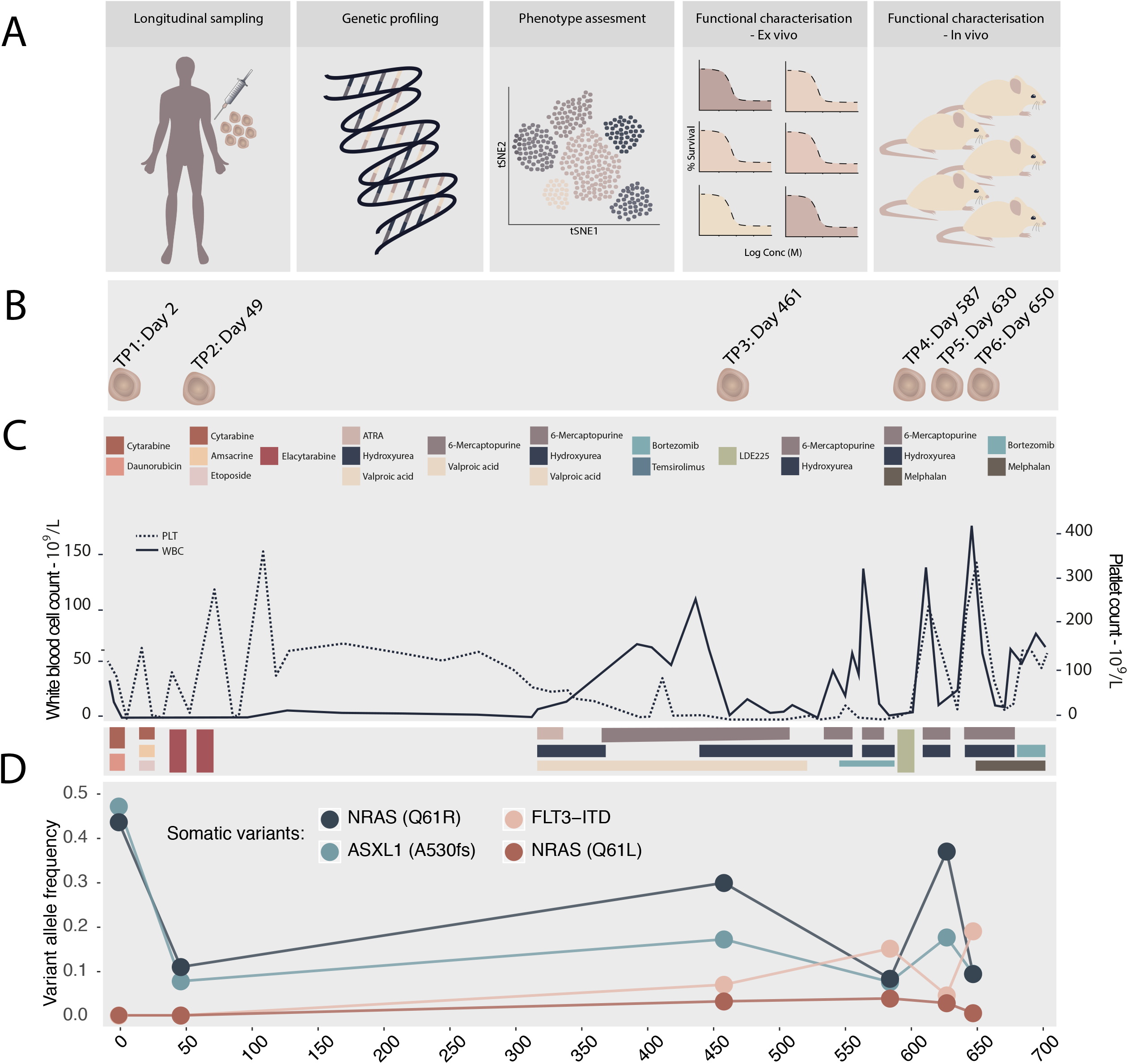
**A**. Concept illustration of the disease characterisation approaches presented in this paper. **B, C** and **D** portrays various aspects of the disease course with a shared x-axis, measuring chronological time in days. **B**. Overview of sampling timepoints of primary material. TP1, TP3, TP5 and TP6 are PB samples. TP2 and TP4 are BM samples. **C**. Outline of the clinical disease course. The left y-axis and whole line represent white blood cell counts while the right y-axis and stippled line denotes platelet counts. Each coloured box denotes an anti-leukaemic medical intervention, as indicated in the bottom panel. **D**. Outline of the AML-related somatic variants and fluctuation of their relative allele distribution over the disease course, as estimated by targeted next generation sequencing. The y-axis represents variant allele frequency (VAF), where 0 means no detected variant allele and 1 means exclusive detection of the variant allele. The figure includes the NRAS Q61L variant, although it was identified below the pre-set threshold of 0.05.

## RESULTS

### Case presentation

The index patient, a male in his late 60’s, presented with fatigue and dyspnoea. The initial diagnostic work up revealed a severe anaemia, thrombocytopenia and concurrent leucocytosis (white blood cell counts 28.3×10^9^/L), suggestive of acute leukaemia. Morphological assessment of a bone marrow (BM) and peripheral blood (PB) smear confirmed the diagnosis, demonstrating 83% and 72% myeloblasts, respectively. Further diagnostic work on day 2 (TP1) by flow cytometry revealed an expanded immature cell population expressing CD34^+^, CD13^+^, CD33^+^, CD117^+^, HLA-DR^+^, MPO^+^. The patient had normal karyotype (46,XY), and FISH analysis was negative. Targeted molecular genetics revealed a *FLT3*-ITD mutation (33 base pairs) and wild-type *NPM1*. The patient classified as intermediate risk in line with ELN2008 and standard induction treatment with cytarabine and daunorubicin commenced on day 4. The disease course is illustrated in Figure 1C. Response assessment at day 24 revealed refractory disease (90 % BM blasts). Subsequently, an induction course with cytarabine, amsacrine and etoposide started on day 25. At re-evaluation of the BM on day 41, 30% blasts remained. The patient was included in a clinical trial and treated with elacytarabine^15^ (ClinicalTrials.gov Identifier: NCT01147939), starting on day 49 (TP2). Complete morphological response with 3% BM blasts was achieved on day 84. Consolidation therapy of elacytarabine was complicated by a severe infection and multiple organ failure. Thereafter he was considered ineligible for an allogeneic haematopoietic stem cell transplant. At 6 months follow-up, the disease had recurred. A disease-stabilising regiment with hydroxyurea, valproic acid and all-trans-retinoic acid according to local protocols^16^ was initiated, starting on day 331. Disease control was not achieved, thus hydroxyurea was exchanged with 6-mercaptopurine. Thereafter, hydroxyurea combined with 6-mercaptopurine^16^, resulting in transient control of peripheral leukocytosis. At genetic re-assessment on day 461 (TP3) the disease was characterised by complex karyotype (Supplementary figure 1). A leukemic sample was analysed by a drug sensitivity and resistance testing (DSRT) pipeline^17^, which indicated sensitivity to the proteasome inhibitor bortezomib. Thus, the patient was treated off-label with four courses of bortezomib and temsirolimus, in line with a multiple myeloma protocol^18^, starting on day 539. The patient was included in an early phase clinical trial on day 587 (TP4), and commenced treatment with the smoothened antagonist sonidegib (LDE225; ClinicalTrials.gov Identifier: NCT01826214). Due to rapid disease progression, the patient was withdrawn from the trial after three weeks and treatment with 6-mercaptopurine and hydroxyurea was reinitiated, resulting in transient disease stabilisation. A new DSRT at day 630 (TP5) suggested acquired sensitivity to melphalan, and the patient was treated with low dose melphalan^19^ in sequence with bortezomib. No objective response was obtained. The patient expired on day 700.

### Temporal fluctuation of genetically defined leukemic cell populations

Longitudinal cytogenetic assessment indicated substantial genetic evolution; from normal to complex karyotype (Supplementary figure 1). The kinetics were further explored by G-band analysis of additional metaphases, array-based copy number analysis and individualised FISH of the samples TP1, 3, 4 and 6 (Supplementary figure 2). At diagnosis (TP1) the leukemic cells were karyotypically normal (46, XY). Copy number analysis revealed a cryptic 2,2 Mb focal deletion of *RUNX1* (del(21)(q22.1q22.1)) at all sample timepoints. Interphase FISH confirmed a *RUNX1* deletion in 80% of the cells at TP1, suggesting an early somatic event. Copy number analysis of TP1 revealed a 0,6Mb focal deletion on chromosome 18 (del(18)(q21.2q21.2)), which extended to 1,7Mb (del(18)(q21.2q21.31)) in samples from TP3, 4 and 6. FISH analysis revealed that 89% of the interphases at TP1 contained the 0,6Mb deletion and 1% the 1,7Mb deletion, whereas at TP3 the relative distribution was 18% and 66%, respectively. G-band analysis of the TP1 sample confirmed a 6q deletion and t(11;18), as seen at TP3, in 3/30 metaphases. In addition to extension of the 18q deletion, TP3 and TP4 were characterised by t(11;18) in all G-banded metaphases, and a focal deletion on chromosome 11 (del(11)(q13.4q13.4)). FISH probes designed towards these regions exposed that the deletions occurred in breakpoints in the translocation between 11q13.4 and 18q21.3. Subsequently, we designed probes verifying a translocation and breakpoints in der(7)t(7;8)(q21.11;q23.2).

**Figure 2.**
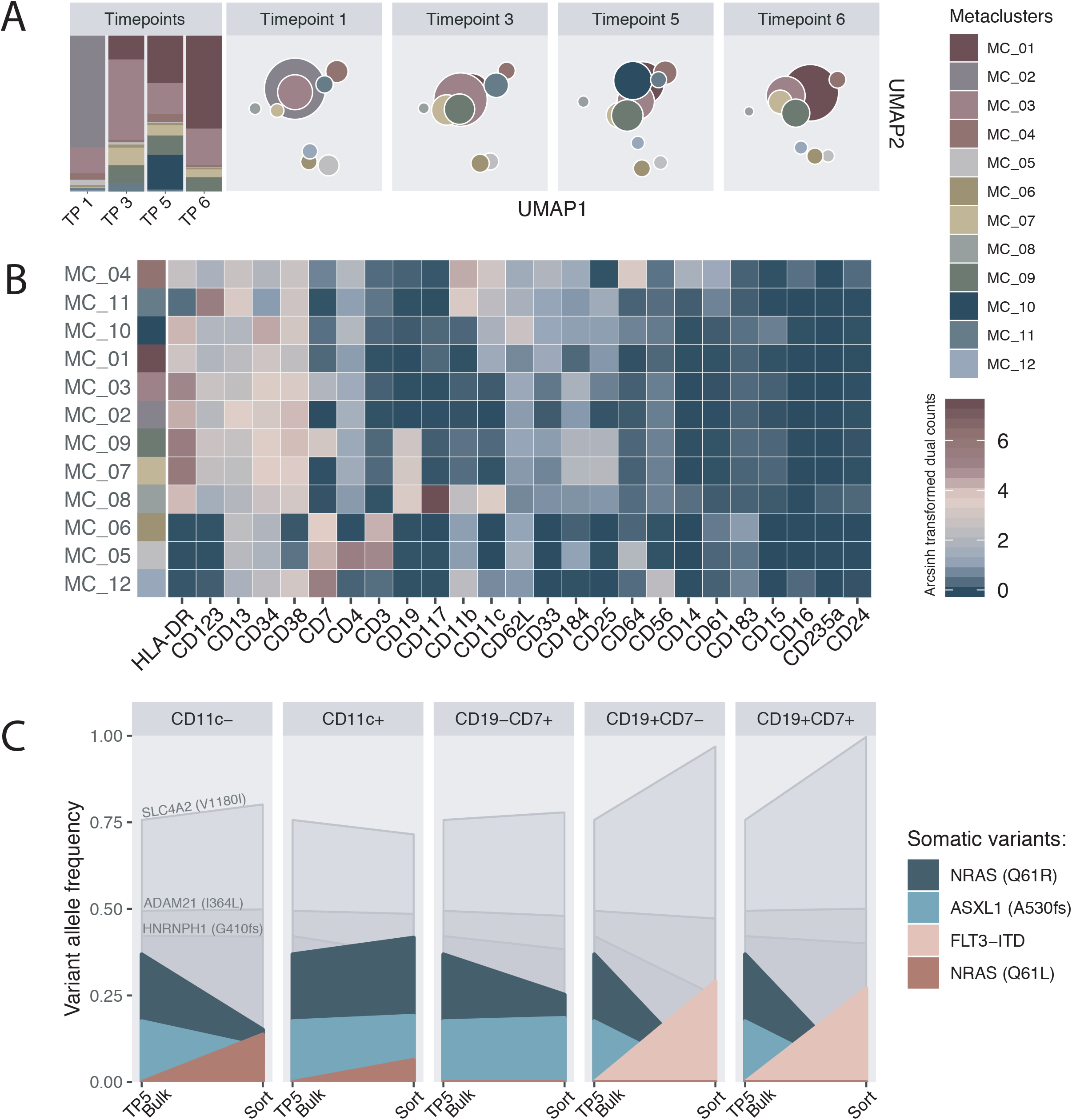
Mass-cytometry data of the primary samples. **A**. Barplot depicting the relative distribution of the 12 metaclusters (MC) per primary sample (n=38. TP1:9, TP3:9, TP5:11, TP6:9). UMAP based on the arcsinh-transformed median dual count of 18 lineage markers. Each node represents a MC within a sample and the nodes are coloured according to respective MC, as annotated in the heatmap in B. The location of the nodes in the map represents the phenotype, based on surface marker expression, while the node size reflects cell numbers. **B**. Heatmap of the median expression of 25 markers. The cells have been gated on CD45+ expression; thus, CD45 is excluded from the figure. Red indicates high and blue indicates low relative expression. **C**. Graphical presentation of the cell sort experiment. Each box represents a sorted cell population. In each box, the y-axis represent VAF and the left x-axis denotes the variant allele distribution in the bulk TP5 sample. The right x-axis indicates the VAF in the sorted sample. The CD11c^+^ samples and the CD11c^-^ are both CD19- and CD7-.

To assess the correlation between molecular arcitechure and changes in karyotype, we re-analysed *FLT3* mutation status and performed fragment analysis on samples from TP1, 3, 4 and 6. At TP1, A low frequency *FLT3*-ITD mutation (ITD/WT ratio=0.03) was detected. At TP3, the ratio had increased to 0.6 and by TP4 two additional ITDs were detected, of an estimated length of 18 and 9 base pairs, respectively. At TP6, the ratio had decreased to 0.2.

Furthermore, whole exome sequencing of samples from TP1, 3, 5 and 6 (all PB) was performed. A total of 36 unique variants in 35 genes were detected, of which nine were shared across all timepoints (Supplementary table 1). However, only 3/36 variants were characterised by a stable variant allele fraction (VAF) above 40% (*SLC4A2* V1180I, *HNRNPH1* G410G, *ADAM21* I364L), while remaining variants fluctuated substantially, suggesting various subpopulations of cells. Two of the 35 genes are recurrently mutated in AML; *ASXL1* and *NRAS*, suggesting involvement in pathogenesis, in addition to the *FLT3*-ITD mutation and the *RUNX1* deletion. Interpreted in concert with copy number analysis and the *FLT3*-ITD ratio, the temporal VAF pattern suggested that the *RUNX1* deletion, *SLC4A2* V1180I, *HNRNPH1* G410G and *ADAM21* I364L were truncal events. *NRAS* Q61R and *FLT3*-ITD, however, seemed to represent independent cell populations, where the *NRAS* Q61R variant was present in cells with the additional variants *ASXL1* A530fs, *FAT4* R1801W, *STIL* S610R, *KCNC2* S630F and *TRAF6* Y418F.

The three trunk mutations, in addition to *FLT3*-ITD, *NRAS* Q61R, *ASXL1* A530fs and *TRAF6* Y418F, were further validated with a capture-based approach. There was concordance between the VAF estimated by whole exome and targeted sequencing (Supplementary figure 4). Additional BM samples obtained at TP2 and TP4 were analysed. Previous reports suggested asymmetric peripheralisation of leukemic cell populations^11^. The addition of BM samples enabled comparison of paired BM and PB samples, where we found high agreement of VAF across the assessed variants (Supplementary figure 5). The VAF pattern of the expanded timeline supported the presence of genetically distinct cell populations: one characterised by *NRAS* Q61R and *ASXL1* A530fs, and one characterised by the *FLT3*-ITD mutation. Further fluctuation of VAF across the remaining timepoints was extensive, suggesting substantial variation of the composition of the leukemic compartment (Figure 1D, Supplementary table 2).

**Figure 3.**
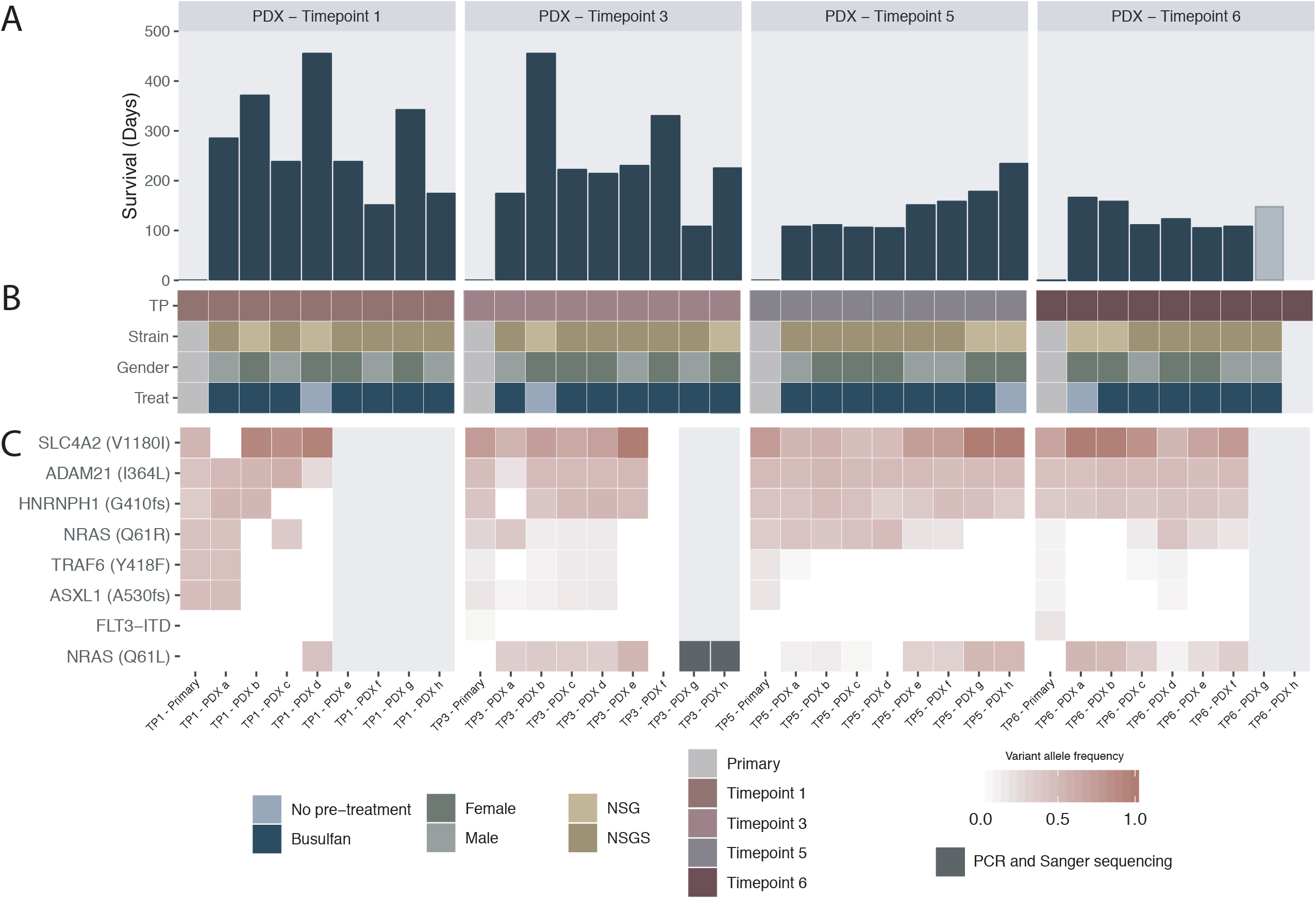
**A**. Survival time (days) of the 31 individual mice included in the PDX experiment, organised by primary sample origin: TP1, TP3, TP5 and TP6, respectively. **B**. Indicators of individual samples including timepoint and mouse strain, sex and myeloablation status. **C**. Heatmap of mutation status of individual samples including the 4 primary samples and the 26 PDX-derived samples of which engraftment was confirmed. Heat indicates VAF. TP1-PDX a and TP6-PDX f are exome sequenced. TP5-PDX h and TP5-PDX a have been characterised by copy number analysis. TP1-PDX e, TP1-PDX f, TP1-PDX g and TP1-PDX h did not engraft. Engraftment of TP3-PDX g, was confirmed by flow cytometry (spleen), mass-cytometry (spleen) and detection of *NRAS* Q61L by Sanger sequencing (BM). Engraftment of TP3-PDX h was confirmed by mass-cytometry (spleen), and detection of *NRAS* Q61L by Sanger sequencing (BM). TP6-PDX g was found dead in the cage and was excluded from all analyses.

**Figure 4.**
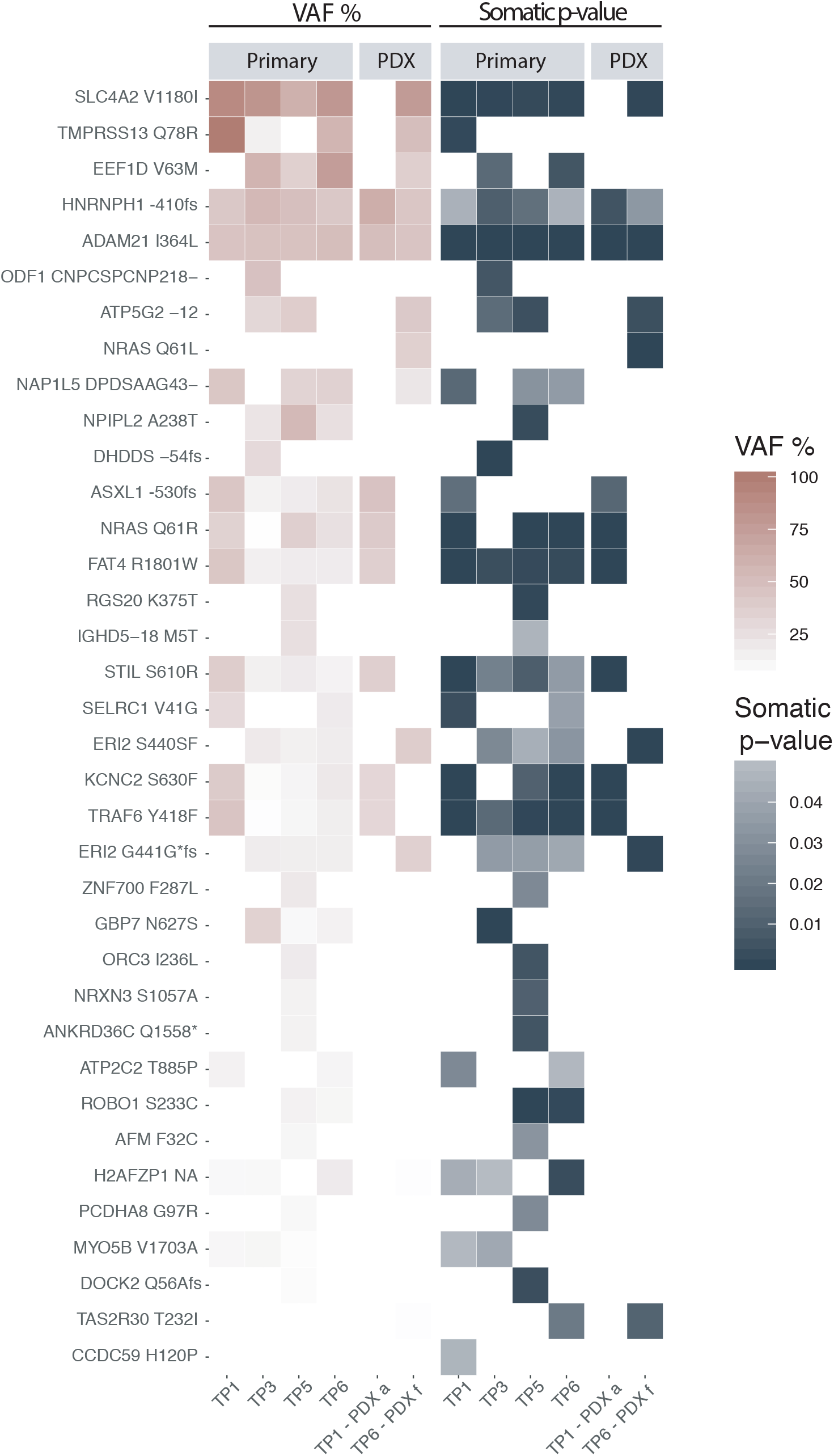
Whole exome sequencing results of the four primary samples (TP1, TP3, TP5 and TP6) and two PDX-derived samples (TP1-PDX a and TP6-PDX f) showing the VAF (left panel) and somatic p-value (right panel).

**Figure 5.**
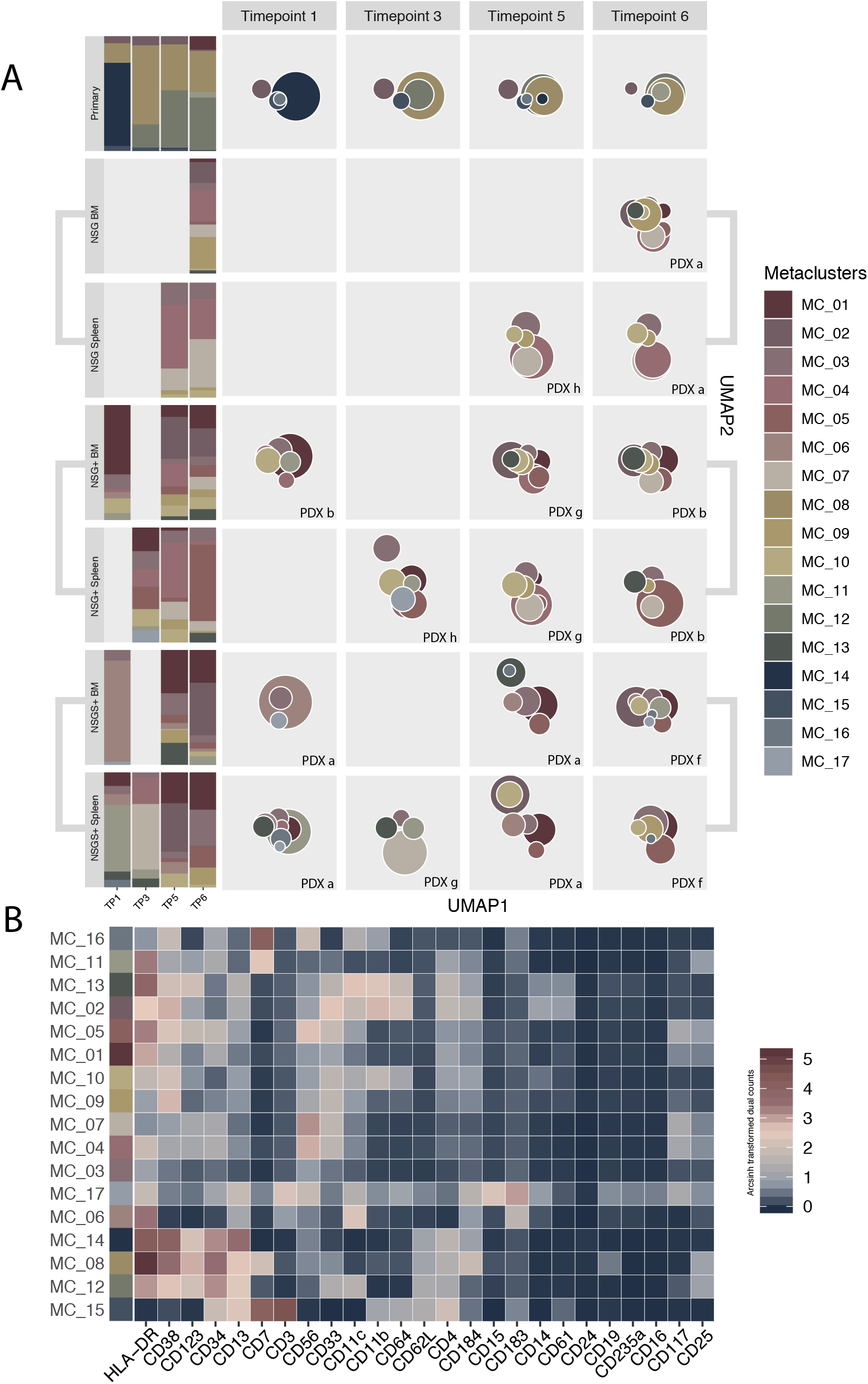
**A**. Outline of the immunophenotypic landscape of the 4 primary samples and 16 PDX samples analysed by mass-cytometry. Each bar in the stacked bar plot and each box in the UMAP plot represents an individual sample, with timepoints organised from left to right and various PDX samples from top to bottom, with the primary samples as reference on top. Intra-sample heterogeneity is indicated by stacked columns to the left where the relative contribution of each MC within individual samples is presented (n=128). In the UMAP plots each node represents a single MC, as identified by the PhenoGraph analysis (K=30). Node proximity correspond to marker expression similarity and the size of each MC indicates cell count. The colour refers to the MC presented in the heatmap in B. **B**. Heatmap of 25/26 markers. Cells are pre-gated for CD45+; hence, this marker is not shown. Colour indicates relative marker expression (arcsinh transformed dual counts, co-factor=5). Red indicates high and blue indicates low relative expression.

### High-dimensional characterisation of immune diversity during treatment

The genomic landscape suggested that the leukaemic samples comprised multiple genetically discrete cell populations. To examine this heterogeneity; we analysed the immunophenotype of samples from TP1, 3, 5 and 6 by mass cytometry with a 26-marker panel. For analysis, a CD45^+^CD235a^-^ gate was explored by a data-driven clustering and meta-clustering approach using Phenograph^20^, resulting in identification of 12 meta-clusters (MCs) (Figure 2A, B and Supplementary table 4). Presumptively normal lymphatic immune subsets clustered in MC06, MC05 and MC12. The remaining MCs were interpreted to represent phenotypic diversity within the leukemic blast compartment. At TP1 the majority of blasts were characterised by the expression of HLA-DR, CD123, CD13, CD34, and CD38 (MC02). At TP3, however, the majority of cells expressed relative higher levels of HLA-DR, CD123 and CD7 and lower levels of the myeloid markers CD13 and CD38 (MC03). In addition, CD184 expression characterised the majority of cells at TP3 (MC03, MC07 and MC09). While the relative frequency of MC03 declined at TP5 and TP6, the relative frequency of MC01, characterised by higher CD11c expression, synchronously increased. TP3, 5 and 6 were further characterised by the expansion and persistence of CD19^+^ cells (MC07, MC08 and MC09).

### Correlation of genotype and immunophenotype

CD19 expression is associated with t(8;21) positive AML^21^. Analogously, CD7 expression is associated with *FLT3*-ITD mutations^22^. The temporal relative frequency pattern of cells expressing CD19 and CD7 resembled the dynamic VAF of *FLT3*-ITD. To deconvolute this relationship, leukaemic cells from TP5 were depleted of CD3 positive cells, live stained and sorted by multi-colour flow cytometry based on surface marker expression of CD7, CD19 and CD11c. This resulted in delineation of five populations: (i). CD19^+^CD7^+^, (ii) CD19^+^CD7^-^, (iii) CD19^-^CD7^+^, (iv) CD19^-^CD7^-^CD11^+^ population, and a dumping channel (v) CD19^-^CD7^-^CD11c^-^. Genomic DNA from the sorted populations was analysed by targeted sequencing, as previously described (Figure 2C). As expected, the various blast populations shared the trunk variants. VAF patterns of the presumed driver mutations, however, varied across the cell subsets (Supplementary table 2). In both CD19^+^ enriched samples, *FLT3*-ITD dominated while the *NRAS* Q61R variant was undetected. Conversely, CD7 was found to be expressed both in *NRAS* Q61R mutated cells and in *FLT3*-ITD positive cells. In the CD19 and CD7 depleted cell populations, a *NRAS* Q61L variant was unexpectedly enriched for. The variant was retrospectively identified in whole exome and targeted sequencing data in TP3, 4, 5 and 6, but with VAF below the filtering threshold. In sum, our results indicate that variation in immunophenotypically defined cell populations in part reflect genetically diverse cell populations.

### Molecular convergence of leukaemia propagating cells

To further investigate the leukemic stem cell compartment, a repopulation assay was performed. We injected primary cells from TP1, 3, 5 and 6 in immunodeficient mice under heterogeneous conditions. The experiment comprised 32 mice; four groups with eight mice per group (Figure 3A and 3B, Supplementary table 5). The groups were composed of two female NSG mice and six NSGS mice of both genders. All but a single NSG mouse in each group were myeloablated with a single dose of busulfan prior to cell injection. Animals were euthanized at humane endpoint. Signs of leukaemic condition developed in 30/30 mice, with a mean latency of 204 days. Human disease engraftment was confirmed by detection of human CD45 cells by flow cytometry in 20/30 mice (Supplementary table 7) and confirmation of patient-specific genetic variants in 26/30 animals (Figure 3C). The experiment demonstrated that all four primary samples could engraft in recipient mice, yet the discrete samples had distinct leukaemia initiating properties and significantly discrepant disease latency. Engraftment was 50% in mice injected with cells from TP1, and 100% in the remaining groups. The samples from TP5 and TP6 had significantly shorter disease latency with a median overall survival of 131 days and 117 days, respectively. In comparison, animals injected with cells from TP1 and TP3 had a median time of disease onset of 328 days and 223 days, respectively (Log rank, p=0.00047) (Figure 3A).

Human cells were extracted from the 26 engrafted specimens where was confirmed. Samples were assessed by PCR, Sanger sequencing and fragment analysis of *NRAS, ASXL1* and *FLT3* (data not shown), followed by targeted sequencing of 24/26 samples (Figure 3C, Supplementary table 2). The trunk mutations were the most recurrent variants identified across the samples, with *ADAM21* I364L found in 23/24 samples, *SLC4A2* V1180I found in 22/24 samples and *HNRNPH1* G410G found in 20/24 samples, respectively. A total of 22/24 samples were NRAS mutated. Across all samples, we identified three genetically defined cellular populations that dominated leukemic initiation: (i) *NRAS* Q61R + *ASXL1* A530fs, (ii) *NRAS* Q61R and (iii) *NRAS* Q61L. 6/22 samples had only the *NRAS* Q61L variant, while 3/22 samples were only *NRAS* Q61R mutated. In the remaining 13 samples, both variants were detected but at distinct frequencies, in support of discrete cell populations. Interestingly, the *NRAS* Q61R variant expanded in 1/8 NSG mice compared with 15/18 NSGS mice. Conversely, the six mice characterised by only the *NRAS* Q61L variant were all of the NSG strain. In 8/16 samples, where a *NRAS* Q61R variant was detected, no *TRAF6* Y418F or *ASXL1* A530fs mutation was identified, strongly suggesting that *NRAS* Q61R preceded these variants. In 4/8 *NRAS* Q61R and *ASXL1* A530fs and/or *TRAF6* Y418F mutated samples, *NRAS* Q61R VAF far exceeded the VAF of *ASXL1* A530fs and/or *TRAF6* Y418F, suggesting the presence of an ancestral *NRAS* Q61R cell population, loss of heterozygosity or both. The same pattern was present in the primary samples TP3 and TP5. Copy number analysis of sample TP5-PDX a revealed dup(1)(q21.1q44) in mosaic which, in part, could account for this pattern, suggesting a 2:1 relationship between the *NRAS* Q61R variant and the germline allele (Supplementary figure 3). *FLT3*-ITD mutated cells contributed to the leukemic disease only in 1/26 mice (TP3-PDX b), where a *FLT3*-ITD of 33 base pairs was detected, however below the cutoff (VAF 0.01). In TP1-PDX b, all three trunk mutations were found but no AML-associated variant was identified. In TP3-PDX f, none of the leukaemia specific variants were detected, although engraftment was confirmed by flow cytometry.

Copy number analysis of TP5-PDX h and TP5-PDX a both revealed deletions corresponding to the karyotype of 46,XY,del(6)(q14.1q22.31),t(11;18)(q21;q22),del(21)(q22.11q22.12) (Supplementary figure 3). The same karyotype was identified in 3/30 metaphases in TP1, suggesting a shared ancestry. To further resolve the ontogeny of the two *NRAS* mutated cell populations, we performed whole exome sequencing of two samples from the PDXs, characterised by the *NRAS* Q61R (TP1-PDX a) and *NRAS* Q61L (TP6-PDX f) variant, respectively (Figure 4, Supplementary table 1). We limited the analysis to high confidence calls in the primary material. The results indicated that only two variants were shared by the two populations: *ADAM21* I364L and *HNRNPH1* G410G. TP1-PDX a shared 8/16 variants identified in TP1, including the *NRAS* Q61R, *ASXL1* A530fs, and *TRAF6* Y418F, but not the *SLC4A2* V1180I variant. In addition, this cell population comprised the variants *FAT4* R1801W, *STIL* S610R, *KCNC2* S630F, supporting that these variants characterise one cell population, as identified in the primary material. TP6-PDX f shared 8/21 variants with the parental TP6 sample. Most were first detected at TP3 and persisted at TP5 and TP6. Based on this pattern we postulate that the *NRAS* Q61L and *FLT3*-ITD mutated populations share a common origin, characterised in part by the mutations identified in TP6-PDX f.

In sum, these results indicated that the leukaemia propagating cells in the mice poorly recapitulated the architecture of the parental samples. Further, the pattern did not correspond to the evolutionary trajectory in the patient. This suggests that the leukaemia initiating properties of the human cells is not merely intrinsic but at least in part defined by contextual conditions.

### Phenotypic relationship between the primary leukemic cells and their PDX-derived progeny

To unravel the phenotypic relationship between primary and PDX samples, a subset of 16 samples from 10 PDXs were analysed by mass cytometry alongside the primary samples. Data from each sample was clustered independently, and meta-clustered using PhenoGraph (Figure 5, Supplementary Figure 6, Supplementary table 6). Hierarchical clustering of the relative MC distribution identified a single sample outlier (TP1-PDX a) and further separated the primary samples from the PDX-expanded samples (Supplementary Figure 6). Although no distinct phenotype was shared by all samples, each phenotype was detected in at least two samples. Extensive immunophenotypic heterogeneity was identified in PDXs derived from the same primary sample. Marked dissimilarities between samples derived from the BM and spleen of the same mouse was observed. Importantly, similarities between the samples appeared more strongly associated with the tissue from which they were harvested than the sample from which they originated. The overall expression profile of CD56 was highly distinct between the PDXs and primary samples, with a frequency of less than 1% of primary cells, compared to a mean fraction of 34.3% of PDX cells.

**Figure 6.**
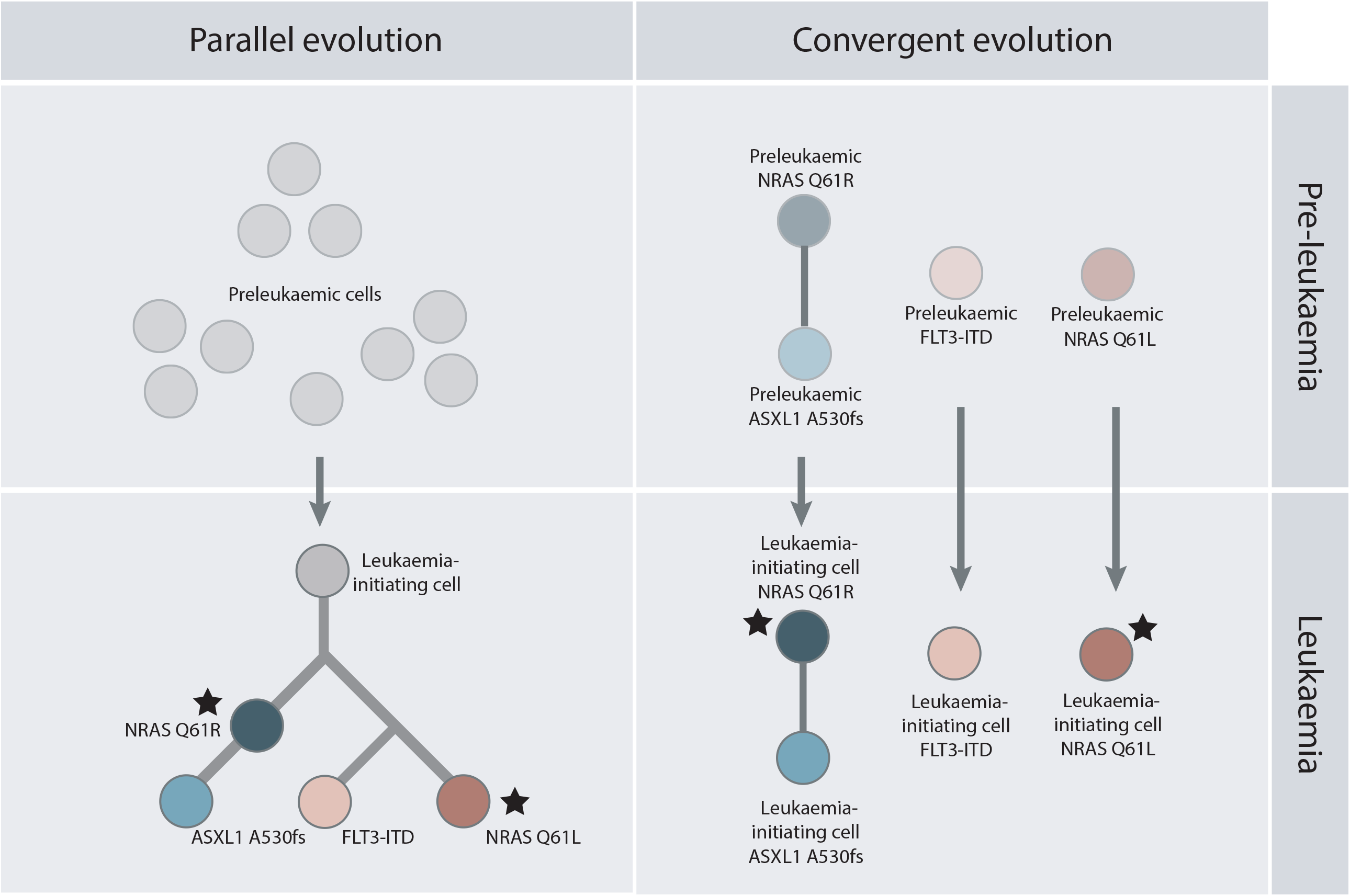
Illustration depicting two alternative trajectories of the leukaemic transition in the index patient. Each circle represents a particular genotype. In evolutionary biology, emergence of similar genotypes or phenotypes across lineages is referred to as parallel or convergent evolution, defined by the relatedness of lineages. Venkatesan and colleagues suggested that a similar terminology can be useful in cancer biology, referring to emerging patterns of similarities descending from one or separate cancer initiating cells^36^. We synthesise that the plurality of independent driver mutations in this patient represent either parallel or convergent evolution. The delineation of a “cancer initiating cell” is instructive in deciding whether this represents a parallel or converging evolutionary pattern. We suggest that the molecular profile characterising the various subpopulations of cells and the estimated timeframe in this patient may indicate plurality of cancer-initiating cells. The clearly converging pattern characterising leukemic propagation in the mice supports this conclusion.

### Functional characterisation through drug sensitivity and resistance testing

We investigated the drug sensitivity of a total of 17 samples, including three primary samples (TP1, 3 and 5) and 14 PDX-derived samples (TP1:0, TP3:5, TP5:4, TP6:4) towards a total of 57 anti-cancer compounds (Supplementary table 8)^23^. The analysis was limited to samples with >70% human CD45+ cells, as assessed by flow cytometry. To obtain a selective DSS (sDSS) score as a metric of the leukaemia-specific effect, the mean DSS score of two healthy BM samples were subtracted from every DSS value. Higher sDSS indicate relative sensitivity and lower values indicate relative resistance. With an arbitrary cut-off set at 5, the majority of compounds were resistant in most samples. Only 220/969 tests were indicative of sensitivity, with great heterogeneity across the samples (Supplementary Figure 7). 19.3% of compounds (11/57) resulted in a sDSS of ≥5 in at least half of the samples (Supplementary table 9). An unsupervised hierarchical clustering analysis resulted in delineation of three clusters separating the TP3-PDX f from the rest. This aligned with the independent ancestry of TP3-PDX f, not characterised by any of the somatic variants that were tested for by targeted sequencing.

## DISCUSSION

Precision haemato-oncology is based on precise categorisation, precise predictions and precise therapeutic targeting. The rationale for the hypothesised superiority of this strategy is based on several assumptions, including: (i) specific and high-resolution molecular understanding can be attained through more research, (ii) causally relevant molecular features are shared by all cells of a leukaemia because of their monoclonal origin, (iii) targeting molecular causal pathways will be efficient, and (iv) provided enough data, precise predictions can be made.

Our results suggest that the description of AML architecture and kinetics is limited by sampling intervals and technological resolution. With each new modality we applied, new levels of complexity emerged, while other findings submerged. Whether our observations are representative for AML patients in general is unknown. However, several lines of evidence, including the work presented here, indicate that disease complexity is far more extensive than characterisation of bulk AML biomaterial suggests. Important examples include the plurality of cell populations characterised by independent driver mutations like *FLT3*-ITD or c-*KIT* at time of diagnosis^24-27^, as well as the rapid emergence of leukemic cell populations characterised by *FLT3* wild-type and alternate driver mutations under FLT3-targeting therapy^28,29^.

It was recently shown through transplantation models of 23 *de novo* AML samples that PDX models in most cases poorly recapitulate the disease in the patients. Further, this study revealed that a high number of these cases indeed harbor multiple leukemic clones at diagnosis^30^. Our longitudinal characterisation of this patient suggests long-term persistence of multiple populations, characterised by discrete patterns of somatic variants, including separate “driver” mutations. Based on VAF patters across primary and PDX samples, we identified at least six distinct populations with independent leukemic potential. This suggests significant genetic heterogeneity within the pool of cells with long-term survival and leukemic propagating properties, frequently assumed to represent leukemic stem cells. By expanding the primary cells in multiple diverse environments, we unraveled a range of alternative evolutionary trajectories, and we further demonstrate that these distinct populations possess distinct phenotypic and functional properties.

Although the various populations all persisted in the patient, their relative contribution to the total disease burden was unequal and fluctuated over time. Evidence suggests that the effect single genetic variants have on competitiveness is not an intrinsic property, but rather a relational function that is influenced by the gene-environment^31-33^. Patterns of convergent molecular evolution suggest that genotypically discrete populations are characterised by distinct intrinsic properties, and that the gene-environments drive these differences. Most striking was the convergent penetrance of the population characterised by *NRAS* Q61L in the NSG mice. Although present and persistent in the patient, this population only represented a minor fraction of the disease.

AML is frequently thought to originate from a single transformed cell^34,35^. This patient was characterised by a low number of public calls shared across all timepoints. A disproportional large number of private somatic variants characterised various leukemic sub-populations. The VAF pattern indicated that the *NRAS* Q61R and *ASXL1* A530fs mutated cell population, that dominated much of the clinical disease course, was further characterised by a minimum of 4 additional exonic non-synonymous variants. The longitudinal distribution of these variants indicated that at least *ASXL1* A530fs and *TRAF6* Y418F occurred after acquisition of the *NRAS* Q61R mutation. Considering the low mutation rate in haematopoietic stem cells^34^ and the relatively stable number of mutations during AML disease courses^10^, we speculate that a shared cellular progeny may have preceded leukemic onset by as much as several decades. Furthermore, co-presentation of *FLT3*-ITD and *NRAS* mutated populations at diagnosis leads us to suspect plurality of leukemic founder clones, implying that the transition between pre-leukaemia and leukaemia may not be categorically monoclonal (Figure 6).

In summary, these observations support that the leukaemic cell environment influences individual leukaemic cells as a selective force. Increased resolution in the characterisation of leukemic samples may reveal additional molecular delineated cells that could contribute in future leukemic trajectories. The question is whether it is feasible to predict or modulate which trajectory will play out. Changes in gene-environment by random acquisition of novel mutations is just the most basic level of unpredictability. Changes in pharmacological pressure, host growth factor responses, endocrine regulation and immune responses may ultimately all contribute in formation of evolutionary trajectories of single cells and cellular ecosystems.

## CONCLUSIONS

Based on converging disease kinetics and engraftment patterns we suggest that leukemogenic transitions may not be categorically monoclonal. Our observations demonstrate that the relationship between leukemic cells is dynamic and relational, and that the trajectory of AML can be influenced by the environment. Thus, we propose that leukemic cell identity may be best conceptualised as a property that can emerge and regress dependent on features of the gene-environment, challenging the conception of leukemic cells as molecular definable entities. This comes into conflict with the monocausal conceptual framework on which precision oncology is largely built, and further suggests limitations with regards to the precision obtainable in molecular categorization, predictive models, and molecular therapeutic targeting.

## Data Availability

All data is available in article supplementary material

## ACKNOWLEDGMENTS

This work was supported by grants provided by The Research Council of Norway, Helse Vest health trust and the Norwegian Cancer Society. The authors are grateful to the patient characterised in this study. We further thank personnel at the Gjertsen-, Bruserud-, McCormack-, Heckman-, and Irish lab for contributing with technical assistance.

## AUTHORSHIP

The study was designed by CE and BTG, and the manuscript composed primarily by CE, MH and BTG. ØB, KP, JI, CH and EMC contributed in experimental design. CE performed experimental work, data analysis, prepared the data for presentation and interpreted the results. AB, EMA and RH contributed with experiments of conventional genetics. SE performed analysis of next generation sequencing experiments. RK, MMM and MH performed drug sensitivity and resistance testing experiments. BF performed mass cytometry experiments, and SEG and MH contributed with mass cytometry data analysis. MH performed flow cytometry experiments. THD and MP contributed in the performance of the PDX experiments. All authors contributed in manuscript preparation.

## METHODS

This study has been performed in accordance with the Declaration of Helsinki and in agreement with approved local protocols. The study was approved by the regional ethics committee; REK Vest, University of Bergen, Bergen, Norway (Project REK2012/2245). Before study initiation, the subject provided written informed consent.

All animal studies were approved by The Norwegian Animal Research Authority and were conducted according to the European Convention for the Protection of Vertebrates Used for Scientific Purposes, in the AAALAC accredited institution – The Laboratory Animal Facility at the University of Bergen.

### Patient material

PB and/or BM samples were sequentially obtained on day 2 (TP1), 49 (TP2), 461 (TP3), 587 (TP4), 630 (TP5), and 650 (TP6) respectively, with day 1 representing the day of initial AML diagnosis (Figure 1B). Once acquired the samples were enriched for mononuclear cells by density gradient centrifugation (Lymphoprep, Axis-Shield, Oslo, Norway), aliquoted and vitally preserved as previously described ^37^. Upon further processing the samples were thawed into 10mL of warm media (90% RPMI 1640 (Sigma-Aldrich Darmstadt, Germany) supplemented with 10% heat inactivated FBS), before they were pelleted by centrifugation at 300g, washed with warm media and pelleted again at 300g before application in various assays. Clinical information was manually curated from the electronic medical records.

### Patient derived xenograft models (PDX)

Female and male NOD-scid IL2rγnull mice (NSG) and NSG™-SGM3 (NSGS) mice, aged 6– 10 weeks old (Vivarium, University of Bergen), were used in the animal experiments. Most mice were myeloablated by a single dose of busulfan (female 25mg/kg, male 15mg/kg) one day prior to initiation of the experiment. On day 0 the mice were injected intravenously (lateral tail vein) with thawed primary cells in a suspension of approximately 1.4×10^6^ - 3.2 ×10^6^ cells in 100 µL sterile NaCl. The samples used included TP1, TP3, TP5 and TP6, which were all mononuclear cells enriched from PB samples. Animals injected with cells from the same sample were distributed across various cages. The animals were housed in groups of up to five individuals in individually ventilated cages. Environmental conditions were kept at a 12-hour dark/night schedule, a constant temperature of 21°C and 50% relative humidity. Bedding, habitat enrichment and cages were autoclaved and changed twice per month. There was a continuous supply of sterile water and food. The general health and condition of the mice was observed daily, and the weight was consistently measured twice weekly. Signs of disease, like dehydration, weight loss, anemia or changes in activity, were assessed by score sheet. Mice found to score sufficiently were euthanized and biological material was harvested. Spleen, femurs and tibia were collected in serum free RPMI media (Sigma-Aldrich) and/or formalin. Femur BM cells and cells from the spleen were washed and prepared in single cell suspension before the samples were cryopreserved and stored at -150 °C suspended in 40% RPMI (Sigma-Aldrich), 50% FBS and 10% DMSO.

### Fluorescence-activated cell sorting (FACS)

10×10^6^ cryopreserved cells from TP5 were thawed and set to rest for 40 minutes in a 5% CO^2^ incubator at 37°C. CD3 positive cells were depleted applying Dynabeads CD3 (Invitrogen) in accordance with producers protocol before the sample was live-stained by standard protocols with the following human antibodies: Mouse Anti-Human CD19, APC (HIB19 RUO, BD Biosciences), Mouse Anti-Human CD7, FITC (M-T701 RUO, BD Biosciences), Mouse Anti-Human CD11c, PE (B-ly6 RUO, BD Biosciences). The cells were sorted on a BD FACSDiva 8.0.1 in two rounds: CD7^+^CD19^+^, CD7^-^CD19^+^, CD7^+^CD19^-^ and a dumping channel. The cells from the dumping channel were then separated by high and low CD11c expression. Collected cells were pelleted by centrifugation and stored at -150 °C before genomic DNA was extracted.

### Flow Cytometry

Cryopreserved cells harvested from PDXs were thawed and either fixed by 2% paraformaldehyde and stained, or live-stained by standard protocols with the following antibodies: Rat Anti-Mouse CD45, FITC (30-F11 RUO, BD Pharmingen) and Mouse Anti-Human CD45, PE-CF594 (HI30 RUO, BD Horizon). Data was acquired on LSR Fortessa (BD Bioscience) and the data was gated and analysed in Cytobank.org (Cytobank Inc.). The fraction of human cells was estimated by Anti-Human CD45/Anti-Mouse CD45 scatterplots, and engraftment was defined as a minimum of 100 cells positively stained by Anti-Human CD45 and comprising > 1% cells positively stained by Anti-Human CD45.

### Time of flight mass cytometry

Mass cytometry analysis was performed as previously described^38^. In brief, the primary cells from TP1, TP3, TP5 and TP6 were thawed and fixed with 2% paraformaldehyde in 1x phosphate buffer and stained with a metal conjugated mass cytometry antibody panel. The primary samples were stained with 26 antibodies, and the PDX samples with 27 antibodies (DVS Sciences, Sunnyvale, CA) as previously published^38^ (Supplementary table 3). Acquisition of the samples was performed on a CyTOF1 mass cytometer (Fluidigm) and the mass cytometry data (.fcs) was pre-processed in Cytobank.org. (Cytobank Inc.). Single cells were identified, and cell debris and doublets removed, by manual gating on Event Length and DNA (Ir-191/193). The single cell data (CD45^+^ CD235a^-^) was arcsinh transformed and clustered using the PhenoGraph algorithm^20^ (k=30) implemented in the cyt graphical interface in MATLAB (R2017b, The Mathworks, Inc.). The data was further processed in R (version 3.5.0)^39^ and R studio (version 1.1.453) and visualised using the uniform manifold approximation and projection (UMAP) algorithm^40^. The following 18 surface markers were used for clustering, meta-clustering and UMAP analysis: CD19, CD117, CD11b, CD4, CD64, CD7, CD34, CD123, CD13, CD62L, CD33, CD11c, CD14, CD38, CD3, HLA-DR, CD184, CD56.

### Drug sensitivity and resistance testing (DSRT)

Drug sensitivity and resistance testing was performed according to the previously published method by Pemovska et al.^17^. Briefly, cryopreserved cells were thawed, incubated with DNase for 1 hour before set to rest for 3 hours in 25% HS-5 conditioned medium (CM)^41^. The cell suspension was subsequently filtered and seeded to pre-drugged 384-well cell culture plates, including each drug in five different concentrations covering a 10 000-fold concentration range, at a cell density of 4 x 10^5^ cells/mL. For the PDX samples, only cells harvested from the spleen were used. After 72 h incubation at 37 °C, 5% CO^2^, cell viability was measured using the CellTiter Glo assay (Promega) and the luminescent signal was quantified with a PHERAstar plate reader (BMG Labtech). Dose response curves were generated based on cell viability readouts with the Dotmatics software (Dotmatics Ltd.) Drug sensitivities were quantified with a drug sensitivity score (DSS), described previously by Yadav *et al* ^23^. A selective DSS (sDSS) score was calculated by subtracting the mean DSS of two healthy cryopreserved BM controls from the individual DSS scores.

### DNA preparation

Isolation of genomic DNA was done either directly from diagnostic samples collected or from cryopreserved cells thawed, as previously described. Samples obtained from the PDXs were depleted for mouse cell contamination by mouse cell depletion kit (MACS Miltenyi Biotec., Sweden) in line with producer guidelines. In animals characterised by splenomegaly, cells harvested from the spleen (n=22) were used. In the remaining cases we used cells harvested from the femurs (n=8). Genomic DNA was isolated either on Qiasymphony with Qiasymphony DNA midi kit or manually with the DNeasy Blood and Tissue kit (Qiagen, Hilden, Germany), in accordance with manufactures protocol.

### G-band analysis, FISH, PCR, Fragment analysis, Sanger sequencing, copy number analysis

Chromosome banding analyses and fluorescence in situ hybridisation (FISH) were performed using standard methods. The following FISH probes where uses: RUNX1RUNX1T1 Dual Colour, dual-fusion, RUNX1 Dual Colour, Tel21q (Abbott Molecular, Des Plaines IL), 15q15.1 split-signal (RP11-103B7, RP11-75C20), 3p13 split-signal (RP11-69BL8, RP11-662021), 111q13.4 split-signal (RP11-101P7, RP11-933G1), 18q21.3q21.31 deletion (RP11-1040L14, RP11-634K3, RP11-691P1, and RP11-353J7) (RP11 probes from Empire Genomics Buffalo NY). Copy number analysis was performed on Applied Biosystems Cytoscan™ HD (Thermo Fisher Scientific, Waltham, MA) following the supplier’s protocols, and analysed in Chromosome Analysis Suite software (version 3.3) with a cut off at 200 kb. Analysis of *FLT3*-ITD was done with fragment analysis and Sanger sequencing as previously described^42^. Mutant allele frequency was calculated using peak area of mutant over wild-type after 25 cycles PCR (Peak Scanner). Mutations in *NRAS* were verified by Sanger sequencing using the Ex3F-ACAAACCAGATAGGCAGAAATG, Ex3R-AACTCTGGTTCCAAGTCATTC primers, 94°C 10min, 35 cycles 94°C 30sec, 58°C 30sec, 72°C 30sec, 72°C 7min, ABI Big Dye™ Terminator v3.1 (all reagents from Thermo Fisher Scientific, Waltham, MA).

### Whole exome sequencing

Whole exome sequencing was performed on the primary samples TP1, TP3, TP5 and TP6 as well as samples derived from two of the PDXs (TP1-PDX a and TP6-PDX f). For filtering of germline variants, we used genomic DNA extracted from a matched skin biopsy acquired at TP3. The experimental and analytic pipeline has previously been described^43^. In short, genomic DNA was amplified and sequencing libraries were produced by exome capture of 3 μg DNA and the NimbleGen SeqCap EZ v2 capture kit (Roche NimbleGen, Madison, WI, USA). Sequencing was performed on a HiSeq 2500 instrument (Illumina, San Diego, CA, USA). Reads were processed and aligned to the human reference genome version GRCh37 using BWA, and somatic variants were called by the VarScan2 somatic algorithm. We limited the analysis to non-synonymous variants, set the strand filter at 1, and required a minimum positional coverage of 8 in the normal sample and a minimum of tumor coverage of 6 reads for inclusion. Annotation was performed by SnpEff 4.03 and Ensembl v68. We set a threshold somatic p-value of ≤0.05 in the primary samples and a minimum VAF of 5%. For all the variants that satisfied the criteria in at least one primary sample, we backtracked and added all identical variants with a higher p-value.

### Targeted sequencing

Selected somatic mutations identified by the whole exome sequencing analysis were validated with a targeted capture-based sequencing approach. The same panel was used for exploratory characterisation of the FACS sorted primary cells and the samples derived from the PDXs. The primers were designed to cover the target sites. Amplicons from different samples were indexed with sample specific index primers so it was possible to pool several samples together. The following overhangs were added to locus specific primers to make them compatible with the index primers: Adapter1 (before locus specific forward primer 5’-3’) ACACTCTTTCCCTACACGACGCTCTTCCGATCT, Adapter2 (before locus specific reverse primer 5’-3’) AGACGTGTGCTCTTCCGATCT. Oligo primer sequences were adapted from Illumina adapter sequences document #1000000002694. Amplicons were amplified with a 2-step PCR protocol. First PCR was done in a volume of 20µL containing 10ng of sample DNA, 10µL of 2× Phusion High-Fidelity PCR Master Mix and 0.375µM of each locus-specific primer. The reaction mixture was brought to a final volume with water. The second PCR was done in a volume of 20µL containing 1µL of the amplified product from the first PCR, 10µL of 2× Phusion High-Fidelity PCR Master Mix, 0.375µM of index primer 1 and 0.375µM of index primer 2. The reaction mixture was brought to the final volume with water. G-Storm GS4 (Somerton) thermal cyclers were used for the PCR reactions. 1^st^ PCR was performed according to the program: initial denaturation at 98°C 30 s, 30 cycles at 98°C for 10 s, at 60°C for 30 s, and at 72°C for 15 s, and the final extension at 72°C for 10 minutes. The second PCR was done according to the program: initial denaturation at 98°C 30 s, 8 cycles at 98°C for 10 s, at 65°C for 30 s, and at 72°C for 20 s, and the final extension at 72°C for 5 minutes. The amplified samples were pooled together without exact quantification. In order to remove dimerized primers, the pool was purified with Agencourt AMPure XP beads (Beckman Coulter, Brea, CA, USA) twice using 0,8x volume of beads compared to the sample pool volume. Purification with 0,8x bead volume removes effectively primer dimers from the sample pool. Agilent 2100 Bioanalyzer (Agilent Genomics, Santa Clara, CA, USA) was used to quantify the amplification performance and yield. Sample pools were sequenced with Illumina MiSeq System using Illumina MiSeq Reagent Kit v2 500 cycles kit (Illumina, San Diego, CA, USA).

### Statistics

The non-parametric Wilcoxon rank sum test was applied for comparison of continuous variables. Survival analyses were performed by generation and comparison of Kaplan-Meier survival curves using the log rank test. A p-value threshold ≤0.05 indicated statistical significance. All statistical analyses were performed in R (version 3.5.0)^39^ and R studio (version 1.1.453) and graphs were made with ggplot2 (version 3.1.0)^44^. Illustrations were made in Adobe illustrator CS6 (version 16.0.0).

