## Supplementary material for "Converging molecular evolution in acute myeloid leukaemia"

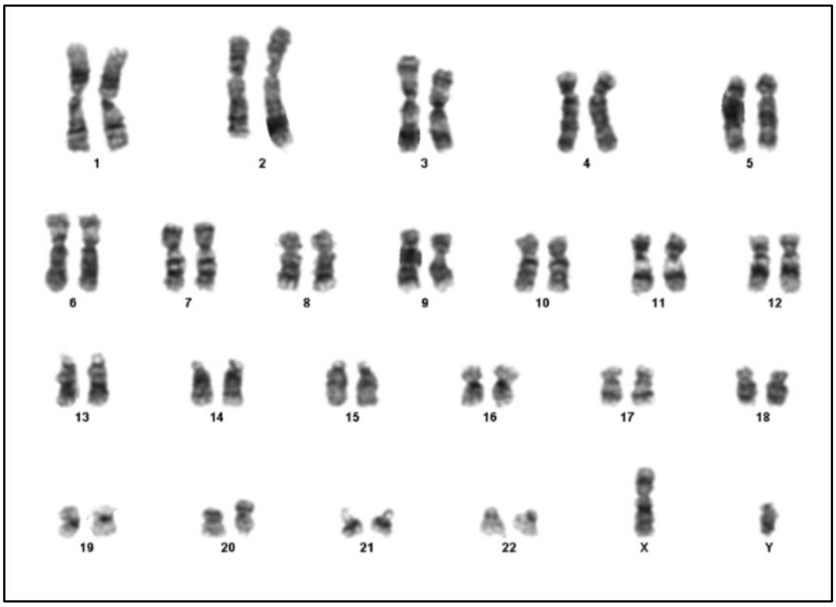


G-band analysis of 20 metaphase after unstimulated cultivation of bone marrow aspirate taken at TP1 show normal karyotype, 46 XY

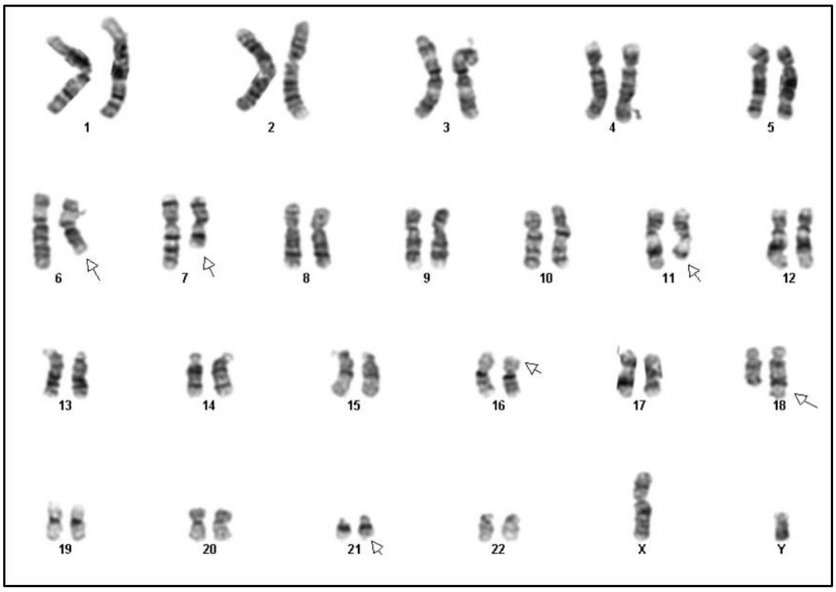


G-band analysis after unstimulated cultivation of bone marrow aspirate taken at TP3 show complex karyotype, 46,XY,del(6)(q16q23),del(7)(q22q35),t(11;18)(q21;q22)[7]/46,idem,t(3;15)(p?21;q?23[3]).

*Supplementary figure 1: G-band analysis of TP1 and TP3.*

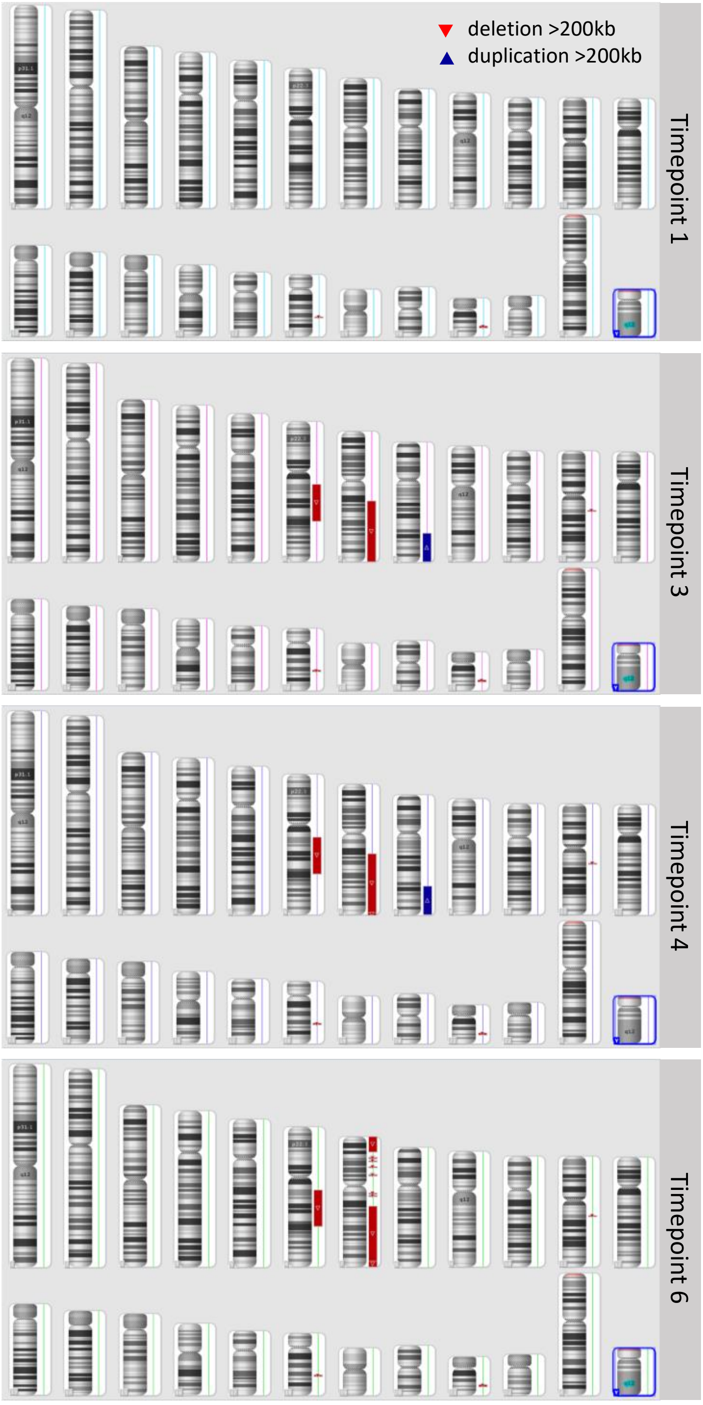


*Supplementary figure 2:Array-based copy number analysis of samples TP1, TP3, TP4 and TP6*

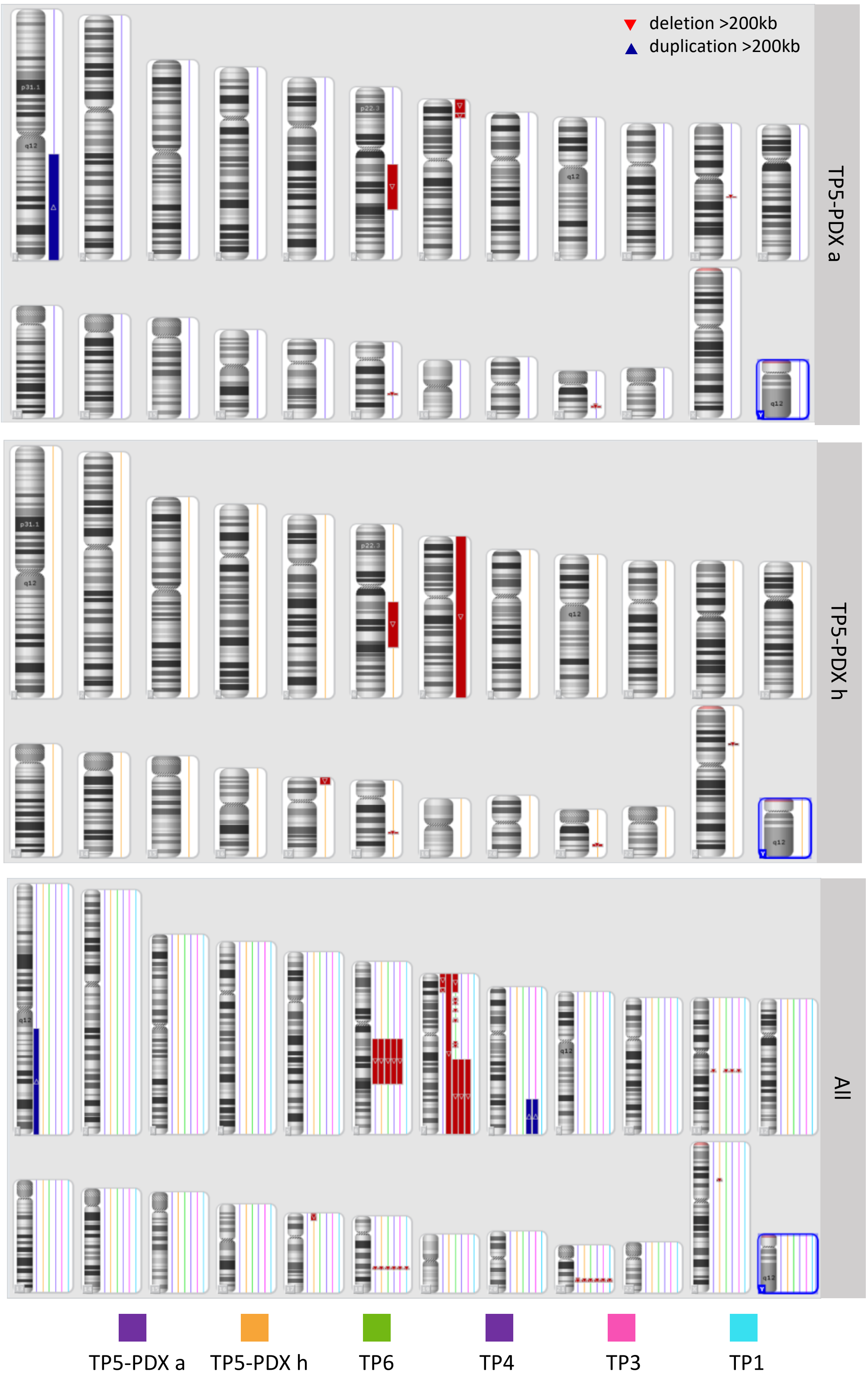


*Supplementary figure 3: Array-based copy number analysis of samples TP5-PDX a, TP5-PDH h, TP1, TP3, TP4 and TP6*

### Supplementary table 1-TP1: Exome sequencing Timepoint 1 – Primary material

| Gene name | Transcript | Chr. | Position | Ref. | Var. | Effect | Amino acid change | Impact | Normal reads 1 | Normal reads 2 | Normal VAF | Tumour reads 1 | Tumour reads 2 | Tumour VAF | Somatic p-value |
|---|---|---|---|---|---|---|---|---|---|---|---|---|---|---|---|
| **ADAM21*** | **ENST00000267499** | **chr14** | **70925306** | **A** | **C** | **NON_SYNONYMOUS_CODING** | **I364L** | **MODERATE** | **27** | **1** | **3.57** | **85** | **70** | **45.16** | **0.00001** |
| **ASXL1*** | **ENST00000375687** | **chr20** | **31021590** | **C** | **CGG** | **FRAME_SHIFT** | **-530?** | **HIGH** | **8** | **0** | **0** | **28** | **22** | **44** | **0.01579** |
| ATP2C2 | ENST00000416219 | chr16 | 84495404 | A | C | NON_SYNONYMOUS_CODING | T885P | MODERATE | 30 | 0 | 0 | 41 | 7 | 14.58 | 0.02787 |
| CCDC59 | ENST00000256151 | chr12 | 82750844 | T | G | NON_SYNONYMOUS_CODING | H120P | MODERATE | 65 | 0 | 0 | 299 | 16 | 5.08 | 0.04648 |
| **FAT4*** | **ENST00000394329** | **chr4** | **126328128** | **C** | **T** | **NON_SYNONYMOUS_CODING** | **R1801W** | **MODERATE** | **36** | **0** | **0** | **90** | **69** | **43.4** | **0.00000** |
| H2AFZP1 | ENST00000416034 | chr21 | 45466387 | G | GT | SPLICE_SITE_ACCEPTOR | NA | HIGH | 37 | 0 | 0 | 95 | 10 | 9.52 | 0.04343 |
| **HNRNPH1*** | **ENST00000329433** | **chr5** | **179043197** | **G** | **GC** | **FRAME_SHIFT** | **-410?** | **HIGH** | **9** | **1** | **10** | **36** | **27** | **42.86** | **0.04507** |
| **KCNC2*** | **ENST00000549446** | **chr12** | **75436913** | **G** | **A** | **NON_SYNONYMOUS_CODING** | **S630F** | **MODERATE** | **74** | **2** | **2.63** | **149** | **100** | **40.16** | **0.00000** |
| MYO5B | ENST00000285039 | chr18 | 47363917 | A | G | NON_SYNONYMOUS_CODING | V1703A | MODERATE | 32 | 0 | 0 | 75 | 9 | 10.71 | 0.04829 |
| NAP1L5 | ENST00000323061 | chr4 | 89618758 | GACCA GCCGC GCTGT CAGGG TC | G | CODON_DELETION | DPDSAAG43- | MODERATE | 9 | 0 | 0 | 18 | 14 | 43.75 | 0.01338 |
| **NRAS*** | **ENST00000369535** | **chr1** | **115256529** | **T** | **C** | **NON_SYNONYMOUS_CODING** | **Q61R** | **MODERATE** | **42** | **0** | **0** | **112** | **59** | **34.5** | **0.00000** |
| SELRC1 | ENST00000371538 | chr1 | 53158524 | A | C | NON_SYNONYMOUS_CODING | V41G | MODERATE | 18 | 0 | 0 | 73 | 31 | 29.81 | 0.00315 |
| **SLC4A2*** | **ENST00000413384** | **chr7** | **150773166** | **G** | **A** | **NON_SYNONYMOUS_CODING** | **V1180I** | **MODERATE** | **8** | **0** | **0** | **1** | **9** | **90** | **0.00021** |
| **STIL*** | **ENST00000371877** | **chr1** | **47746300** | **A** | **T** | **NON_SYNONYMOUS_CODING** | **S610R** | **MODERATE** | **25** | **0** | **0** | **91** | **58** | **38.93** | **0.00001** |
| TMPRSS13 | ENST00000445164 | chr11 | 117789342 | T | C | NON_SYNONYMOUS_CODING | Q78R | MODERATE | 8 | 1 | 11.11 | 0 | 6 | 100 | 0.00140 |
| **TRAF6*** | **ENST00000526995** | **chr11** | **36511704** | **T** | **A** | **NON_SYNONYMOUS_CODING** | **Y418F** | **MODERATE** | **76** | **0** | **0** | **143** | **116** | **44.79** | **0.00000** |

*Mutations shared across all primary samples

#### Supplementary table 1-TP3: Exome sequencing Timepoint 3 - Primary material

| Gene name | Transcript | Chr. | Position | Ref. | Var. | Effect | Amino acid change | Impact | Normal reads 1 | Normal reads 2 | Normal VAF | Tumour reads 1 | Tumour reads 2 | Tumour VAF | Somatic p-value |
|---|---|---|---|---|---|---|---|---|---|---|---|---|---|---|---|
| **ADAM21*** | **ENST00000267499** | **chr14** | **70925306** | **A** | **C** | **NON_SYNONYMOUS_CODING** | **I364L** | **MODERATE** | **27** | **1** | **3.57** | **70** | **63** | **47.37** | **0.00000** |
| **ASXL1*** | **ENST00000375687** | **chr20** | **31021590** | **C** | **CGG** | **FRAME_SHIFT** | **-530?** | **HIGH** | **8** | **0** | **0** | **55** | **9** | **14.06** | **0.32358** |
| ATP5G2 | ENST00000394349 | chr12 | 54069941 | TAG | T | FRAME_SHIFT | -12 | HIGH | 13 | 0 | 0 | 40 | 18 | 31.03 | 0.01448 |
| DHDDS | ENST00000360009 | chr1 | 26764756 | G | GCC | FRAME_SHIFT | -54? | HIGH | 34 | 1 | 2.86 | 118 | 49 | 29.34 | 0.00025 |
| EEF1D | ENST00000532741 | chr8 | 144672215 | C | T | NON_SYNONYMOUS_CODING | V63M | MODERATE | 8 | 1 | 11.11 | 17 | 23 | 57.5 | 0.01363 |
| ERI2 | ENST00000357967 | chr16 | 20809799 | T | TCTAC | FRAME_SHIFT | -441V? | HIGH | 20 | 0 | 0 | 67 | 14 | 17.28 | 0.03554 |
| ERI2 | ENST00000357967 | chr16 | 20809802 | A | AAAT | CODON_CHANGE_PLUS_CODON_INSERTION | S440YS | MODERATE | 20 | 0 | 0 | 61 | 14 | 18.67 | 0.02743 |
| **FAT4*** | **ENST00000394329** | **chr4** | **126328128** | **C** | **T** | **NON_SYNONYMOUS_CODING** | **R1801W** | **MODERATE** | **36** | **0** | **0** | **157** | **30** | **16.04** | **0.00335** |
| GBP7 | ENST00000294671 | chr1 | 89597869 | T | C | NON_SYNONYMOUS_CODING | N627S | MODERATE | 31 | 1 | 3.12 | 139 | 74 | 34.74 | 0.00006 |
| H2AFZP1 | ENST00000416034 | chr21 | 45466387 | G | GT | SPLICE_SITE_ACCEPTOR | NA | HIGH | 37 | 0 | 0 | 88 | 9 | 9.28 | 0.04899 |
| **HNRNPH1*** | **ENST00000329433** | **chr5** | **179043197** | **G** | **GC** | **FRAME_SHIFT** | **-410?** | **HIGH** | **9** | **1** | **10** | **37** | **43** | **53.75** | **0.00919** |
| **KCNC2*** | **ENST00000549446** | **chr12** | **75436913** | **G** | **A** | **NON_SYNONYMOUS_CODING** | **S630F** | **MODERATE** | **74** | **2** | **2.63** | **202** | **18** | **8.18** | **0.07380** |
| MYO5B | ENST00000285039 | chr18 | 47363917 | A | G | NON_SYNONYMOUS_CODING | V1703A | MODERATE | 32 | 0 | 0 | 61 | 8 | 11.59 | 0.04137 |
| NPIPL2 | ENST00000429990 | chr16 | 74425358 | G | A | NON_SYNONYMOUS_CODING | A238T | MODERATE | 10 | 0 | 0 | 18 | 5 | 21.74 | 0.14178 |
| **NRAS*** | **ENST00000369535** | **chr1** | **115256529** | **T** | **C** | **NON_SYNONYMOUS_CODING** | **Q61R** | **MODERATE** | **42** | **0** | **0** | **173** | **10** | **5.46** | **0.12081** |
| ODF1 | ENST00000285402 | chr8 | 103573010 | CTGCAACCCCTGCAGCCCCTGCAACCCG | C | CODON_DELETION | CNPCSPCNP218- | MODERATE | 9 | 0 | 0 | 29 | 26 | 47.27 | 0.00592 |
| **SLC4A2*** | **ENST00000413384** | **chr7** | **150773166** | **G** | **A** | **NON_SYNONYMOUS_CODING** | **V1180I** | **MODERATE** | **8** | **0** | **0** | **2** | **9** | **81.82** | **0.00060** |
| **STIL*** | **ENST00000371877** | **chr1** | **47746300** | **A** | **T** | **NON_SYNONYMOUS_CODING** | **S610R** | **MODERATE** | **25** | **0** | **0** | **111** | **20** | **15.27** | **0.02354** |
| TMPRSS13 | ENST00000445164 | chr11 | 117789342 | T | C | NON_SYNONYMOUS_CODING | Q78R | MODERATE | 8 | 1 | 11.11 | 17 | 3 | 15 | 0.63597 |
| **TRAF6*** | **ENST00000526995** | **chr11** | **36511704** | **T** | **A** | **NON_SYNONYMOUS_CODING** | **Y418F** | **MODERATE** | **76** | **0** | **0** | **239** | **16** | **6.27** | **0.01376** |

*Mutations shared across all primary samples

#### Supplementary table 1-TP5: Exome sequencing Timepoint 5 - Primary material

| Gene name | Transcript | Chr. | Position | Ref. | Var. | Effect | Amino acid change | Impact | Normal reads 1 | Normal reads 2 | Normal VAF | Tumour reads 1 | Tumour reads 2 | Tumour VAF | Somatic p-value |
|---|---|---|---|---|---|---|---|---|---|---|---|---|---|---|---|
| **ADAM21*** | **ENST00000267499** | **chr14** | **70925306** | **A** | **C** | **NON_SYNONYMOUS_CODING** | **I364L** | **MODERATE** | **27** | **1** | **3.57** | **94** | **85** | **47.49** | **0.00000** |
| AFM | ENST00000226355 | chr4 | 74349664 | T | G | NON_SYNONYMOUS_CODING | F32C | MODERATE | 31 | 0 | 0 | 198 | 25 | 11.21 | 0.03237 |
| ANKRD36C | ENST00000456556 | chr2 | 96521337 | G | A | STOP_GAINED | Q1558* | HIGH | 87 | 4 | 4.4 | 400 | 65 | 13.98 | 0.00523 |
| **ASXL1*** | **ENST00000421155** | **chr20** | **31021590** | **C** | **CGG** | **FRAME_SHIFT** | **A530A?** | **HIGH** | **8** | **0** | **0** | **60** | **13** | **17.81** | **0.22982** |
| ATP5G2 | ENST00000394349 | chr12 | 54069941 | TAG | T | FRAME_SHIFT | -12 | HIGH | 13 | 0 | 0 | 38 | 24 | 38.71 | 0.00376 |
| DOCK2 | ENST00000256935 | chr5 | 169096329 | A | AG | FRAME_SHIFT | Q56A? | HIGH | 82 | 0 | 0 | 370 | 30 | 7.5 | 0.00306 |
| EEF1D | ENST00000532741 | chr8 | 144672215 | C | T | NON_SYNONYMOUS_CODING | V63M | MODERATE | 8 | 1 | 11.11 | 14 | 8 | 36.36 | 0.16743 |
| ERI2 | ENST00000357967 | chr16 | 20809799 | T | TCTAC | FRAME_SHIFT | G441G*? | HIGH | 20 | 0 | 0 | 123 | 24 | 16.33 | 0.03645 |
| ERI2 | ENST00000357967 | chr16 | 20809802 | A | AAAT | CODON_CHANGE_PLUS_ CODON_INSERTION | S440SF | MODERATE | 20 | 0 | 0 | 126 | 23 | 15.44 | 0.04419 |
| **FAT4*** | **ENST00000394329** | **chr4** | **126328128** | **C** | **T** | **NON_SYNONYMOUS_CODING** | **R1801W** | **MODERATE** | **36** | **0** | **0** | **147** | **32** | **17.88** | **0.00169** |
| GBP7 | ENST00000294671 | chr1 | 89597869 | T | C | NON_SYNONYMOUS_CODING | N627S | MODERATE | 31 | 1 | 3.12 | 187 | 20 | 9.66 | 0.19421 |
| **HNRNPH1*** | **ENST00000329433** | **chr5** | **179043197** | **G** | **GC** | **FRAME_SHIFT** | **G410G?** | **HIGH** | **9** | **1** | **10** | **73** | **70** | **48.95** | **0.01600** |
| IGHD5-18 | ENST00000390575 | chr14 | 106359406 | A | G | NON_SYNONYMOUS_CODING | M5T | MODERATE | 18 | 1 | 5.26 | 56 | 19 | 25.33 | 0.04628 |
| **KCNC2*** | **ENST00000549446** | **chr12** | **75436913** | **G** | **A** | **NON_SYNONYMOUS_CODING** | **S630F** | **MODERATE** | **74** | **2** | **2.63** | **167** | **23** | **12.11** | **0.01012** |
| MYO5B | ENST00000285039 | chr18 | 47363917 | A | G | NON_SYNONYMOUS_CODING | V1703A | MODERATE | 32 | 0 | 0 | 85 | 6 | 6.59 | 0.15686 |
| NAP1L5 | ENST00000323061 | chr4 | 89618758 | GACCA GCCGC GCTGT CAGGG TC | G | CODON_DELETION | DPDSAAG43 - | MODERATE | 9 | 0 | 0 | 55 | 28 | 33.73 | 0.03170 |
| NPIPL2 | ENST00000429990 | chr16 | 74425358 | G | A | NON_SYNONYMOUS_CODING | A238T | MODERATE | 10 | 0 | 0 | 14 | 16 | 53.33 | 0.00231 |
| **NRAS*** | **ENST00000369535** | **chr1** | **115256529** | **T** | **C** | **NON_SYNONYMOUS_CODING** | **Q61R** | **MODERATE** | **42** | **0** | **0** | **93** | **55** | **37.16** | **0.00000** |
| NRXN3 | ENST00000330071 | chr14 | 79454391 | T | G | NON_SYNONYMOUS_CODING | S1057A | MODERATE | 40 | 0 | 0 | 55 | 9 | 14.06 | 0.01002 |
| ORC3 | ENST00000257789 | chr6 | 88318940 | A | C | NON_SYNONYMOUS_CODING | I236L | MODERATE | 32 | 0 | 0 | 63 | 14 | 18.18 | 0.00524 |
| PCDHA8 | ENST00000531613 | chr5 | 140221195 | G | C | NON_SYNONYMOUS_CODING | G97R | MODERATE | 38 | 0 | 0 | 164 | 18 | 9.89 | 0.02823 |
| RGS20 | ENST00000297313 | chr8 | 54870975 | A | C | NON_SYNONYMOUS_CODING | K375T | MODERATE | 32 | 0 | 0 | 41 | 14 | 25.45 | 0.00080 |
| ROBO1 | ENST00000398414 | chr3 | 78766994 | T | A | NON_SYNONYMOUS_CODING | S233C | MODERATE | 60 | 0 | 0 | 203 | 35 | 14.71 | 0.00022 |
| **SLC4A2*** | **ENST00000413384** | **chr7** | **150773166** | **G** | **A** | **NON_SYNONYMOUS_CODING** | **V1180I** | **MODERATE** | **8** | **0** | **0** | **16** | **25** | **60.98** | **0.00163** |
| **STIL*** | **ENST00000371877** | **chr1** | **47746300** | **A** | **T** | **NON_SYNONYMOUS_CODING** | **S610R** | **MODERATE** | **25** | **0** | **0** | **195** | **43** | **18.07** | **0.00907** |
| **TRAF6*** | **ENST00000526995** | **chr11** | **36511704** | **T** | **A** | **NON_SYNONYMOUS_CODING** | **Y418F** | **MODERATE** | **76** | **0** | **0** | **201** | **25** | **11.06** | **0.00050** |
| ZNF700 | ENST00000254321 | chr19 | 12059700 | T | G | NON_SYNONYMOUS_CODING | F287L | MODERATE | 18 | 0 | 0 | 68 | 17 | 20 | 0.02779 |

*Mutations shared across all primary samples

#### Supplementary table 1-TP6: Exome sequencing Timepoint 6 - Primary material

| Gene name | Transcript | Chr. | Position | Ref. | Var. | Effect | Amino acid change | Impact | Normal reads 1 | Normal reads 2 | Normal VAF | Tumour reads 1 | Tumour reads 2 | Tumour VAF | Somatic p-value |
|---|---|---|---|---|---|---|---|---|---|---|---|---|---|---|---|
| **ADAM21*** | **ENST00000267499** | **chr14** | **70925306** | **A** | **C** | **NON_SYNONYMOUS_CODING** | **I364L** | **MODERATE** | **27** | **1** | **3.57** | **60** | **61** | **50.41** | **0.00000** |
| **ASXL1*** | **ENST00000375687** | **chr20** | **31021590** | **C** | **CGG** | **FRAME_SHIFT** | **-530?** | **HIGH** | **8** | **0** | **0** | **30** | **9** | **23.08** | **0.15552** |
| ATP2C2 | ENST00000416219 | chr16 | 84495404 | A | C | NON_SYNONYMOUS_CODING | T885P | MODERATE | 30 | 0 | 0 | 51 | 7 | 12.07 | 0.04736 |
| EEF1D | ENST00000532741 | chr8 | 144672215 | C | T | NON_SYNONYMOUS_CODING | V63M | MODERATE | 8 | 1 | 11.11 | 3 | 9 | 75 | 0.00580 |
| ERI2 | ENST00000357967 | chr16 | 20809799 | T | TCTAC | FRAME_SHIFT | -441V? | HIGH | 20 | 0 | 0 | 88 | 17 | 16.19 | 0.04102 |
| ERI2 | ENST00000357967 | chr16 | 20809802 | A | AAAT | CODON_CHANGE_PLUS_CODON_INSERTION | S440YS | MODERATE | 20 | 0 | 0 | 81 | 17 | 17.35 | 0.03272 |
| **FAT4*** | **ENST00000394329** | **chr4** | **126328128** | **C** | **T** | **NON_SYNONYMOUS_CODING** | **R1801W** | **MODERATE** | **36** | **0** | **0** | **115** | **25** | **17.86** | **0.00201** |
| GBP7 | ENST00000294671 | chr1 | 89597869 | T | C | NON_SYNONYMOUS_CODING | N627S | MODERATE | 31 | 1 | 3.12 | 175 | 30 | 14.63 | 0.05302 |
| H2AFZP1 | ENST00000416034 | chr21 | 45466387 | G | GT | SPLICE_SITE_ACCEPTOR | NA | HIGH | 37 | 0 | 0 | 63 | 14 | 18.18 | 0.00269 |
| **HNRNPH1*** | **ENST00000329433** | **chr5** | **179043197** | **G** | **GC** | **FRAME_SHIFT** | **-410?** | **HIGH** | **9** | **1** | **10** | **45** | **33** | **42.31** | **0.04536** |
| **KCNC2*** | **ENST00000549446** | **chr12** | **75436913** | **G** | **A** | **NON_SYNONYMOUS_CODING** | **S630F** | **MODERATE** | **74** | **2** | **2.63** | **204** | **51** | **20** | **0.00005** |
| NAP1L5 | ENST00000323061 | chr4 | 89618758 | GACCAGCCGCGCTGTCAGGGTC | G | CODON_DELETION | DPDSAAG43- | MODERATE | 9 | 0 | 0 | 22 | 12 | 35.29 | 0.03575 |
| NPIPL2 | ENST00000429990 | chr16 | 74425358 | G | A | NON_SYNONYMOUS_CODING | A238T | MODERATE | 10 | 0 | 0 | 24 | 8 | 25 | 0.08912 |
| **NRAS*** | **ENST00000369535** | **chr1** | **115256529** | **T** | **C** | **NON_SYNONYMOUS_CODING** | **Q61R** | **MODERATE** | **42** | **0** | **0** | **121** | **40** | **24.84** | **0.00003** |
| ROBO1 | ENST00000398414 | chr3 | 78766994 | T | A | NON_SYNONYMOUS_CODING | S233C | MODERATE | 60 | 0 | 0 | 178 | 24 | 11.88 | 0.00139 |
| SELRC1 | ENST00000371538 | chr1 | 53158524 | A | C | NON_SYNONYMOUS_CODING | V41G | MODERATE | 18 | 0 | 0 | 76 | 17 | 18.28 | 0.03784 |
| **SLC4A2*** | **ENST00000413384** | **chr7** | **150773166** | **G** | **A** | **NON_SYNONYMOUS_CODING** | **V1180I** | **MODERATE** | **8** | **0** | **0** | **2** | **8** | **80** | **0.00103** |
| **STIL*** | **ENST00000371877** | **chr1** | **47746300** | **A** | **T** | **NON_SYNONYMOUS_CODING** | **S610R** | **MODERATE** | **25** | **0** | **0** | **113** | **18** | **13.74** | **0.03516** |
| TAS2R30 | ENST00000539585 | chr12 | 11286149 | G | A | NON_SYNONYMOUS_CODING | T232I | MODERATE | 86 | 0 | 0 | 240 | 13 | 5.14 | 0.02056 |
| TMPRSS13 | ENST00000445164 | chr11 | 117789342 | T | C | NON_SYNONYMOUS_CODING | Q78R | MODERATE | 8 | 1 | 11.11 | 3 | 4 | 57.14 | 0.07692 |
| **TRAF6*** | **ENST00000526995** | **chr11** | **36511704** | **T** | **A** | **NON_SYNONYMOUS_CODING** | **Y418F** | **MODERATE** | **76** | **0** | **0** | **209** | **42** | **16.73** | **0.00001** |

*Mutations shared across all primary samples

#### Supplementary table 1-TP1-PDX a: Exome sequencing Timepoint 1 - PDX a

| Gene name | Transcript | Chr. | Position | Ref. | Var. | Effect | Amino acid change | Impact | Normal reads 1 | Normal reads 2 | Normal VAF | Tumour reads 1 | Tumour reads 2 | Tumour VAF | Somatic p-value |
|---|---|---|---|---|---|---|---|---|---|---|---|---|---|---|---|
| ADAM21 | ENST00000267499 | chr14 | 70925306 | A | C | NON_SYNONYMOUS_CODING | I364L | MODERATE | 27 | 1 | 3.57 | 14 | 14 | 50 | 0.00007 |
| ASXL1 | ENST00000421155 | chr20 | 31021590 | C | CGG | FRAME_SHIFT | A530A? | HIGH | 8 | 0 | 0 | 27 | 23 | 46 | 0.01228 |
| FAT4 | ENST00000394329 | chr4 | 126328128 | C | T | NON_SYNONYMOUS_CODING | R1801W | MODERATE | 36 | 0 | 0 | 56 | 33 | 37.08 | 0.00000 |
| HNRNPH1 | ENST00000329433 | chr5 | 179043197 | G | GC | FRAME_SHIFT | G410G? | HIGH | 9 | 1 | 10 | 10 | 17 | 62.96 | 0.00504 |
| KCNC2 | ENST00000549446 | chr12 | 75436913 | G | A | NON_SYNONYMOUS_CODING | S630F | MODERATE | 74 | 2 | 2.63 | 53 | 25 | 32.05 | 0.00000 |
| NRAS | ENST00000369535 | chr1 | 115256529 | T | C | NON_SYNONYMOUS_CODING | Q61R | MODERATE | 42 | 0 | 0 | 58 | 40 | 40.82 | 0.00000 |
| STIL | ENST00000371877 | chr1 | 47746300 | A | T | NON_SYNONYMOUS_CODING | S610R | MODERATE | 25 | 0 | 0 | 43 | 25 | 36.76 | 0.00009 |
| TRAF6 | ENST00000526995 | chr11 | 36511704 | T | A | NON_SYNONYMOUS_CODING | Y418F | MODERATE | 76 | 0 | 0 | 100 | 47 | 31.97 | 0.00000 |

#### Supplementary table 1-TP6-PDX f: Exome sequencing Timepoint 6 - PDX f

| Gene name | Transcript | Chr. | Position | Ref. | Var. | Effect | Amino acid change | Impact | Normal reads 1 | Normal reads 2 | Normal VAF | Tumour reads 1 | Tumour reads 2 | Tumour VAF | Somatic p-value |
|---|---|---|---|---|---|---|---|---|---|---|---|---|---|---|---|
| ATP5G2 | ENST00000394349 | chr12 | 54069941.00 | TAG | T | FRAME_SHIFT | -12 | HIGH | 13 | 0 | 0 | 23 | 16 | 41.03 | 0.00364 |
| EEF1D | ENST00000532741 | chr8 | 144672215.00 | C | T | NON_SYNONYMOUS_CODING | V63M | MODERATE | 8 | 1 | 11.11 | 10 | 6 | 37.5 | 0.17373 |
| ERI2 | ENST00000357967 | chr16 | 20809799.00 | T | TCTAC | FRAME_SHIFT | G441G*? | HIGH | 20 | 0 | 0 | 58 | 32 | 35.56 | 0.00045 |
| ERI2 | ENST00000357967 | chr16 | 20809802.00 | A | AAAT | CODON_CHANGE_PLUS_CODON_INSERTION | S440SF | MODERATE | 20 | 0 | 0 | 50 | 32 | 39.02 | 0.00019 |
| H2AFZP1 | ENST00000416034 | chr21 | 45466387.00 | G | GT | SPLICE_SITE_ACCEPTOR | NA | HIGH | 37 | 0 | 0 | 201 | 13 | 6.07 | 0.11896 |
| HNRNPH1 | ENST00000329433 | chr5 | 179043197.00 | G | GC | FRAME_SHIFT | G410G? | HIGH | 9 | 1 | 10 | 53 | 42 | 44.21 | 0.03397 |
| NAP1L5 | ENST00000323061 | chr4 | 89618758.00 | GACCAGCCGCGCTGTCAGGGTC | G | CODON_DELETION | DPDSAAG43- | MODERATE | 9 | 0 | 0 | 60 | 16 | 21.05 | 0.13764 |
| NRAS | ENST00000369535 | chr1 | 115256529.00 | T | A | NON_SYNONYMOUS_CODING | Q61L | MODERATE | 42 | 0 | 0 | 103 | 58 | 36.02 | 0.00000 |
| SLC4A2 | ENST00000413384 | chr7 | 150773166.00 | G | A | NON_SYNONYMOUS_CODING | V1180I | MODERATE | 8 | 0 | 0 | 5 | 16 | 76.19 | 0.00030 |
| TAS2R30 | ENST00000539585 | chr12 | 11286149.00 | G | A | NON_SYNONYMOUS_CODING | T232I | MODERATE | 86 | 0 | 0 | 329 | 20 | 5.73 | 0.01092 |
| TMPRSS13 | ENST00000445164 | chr11 | 117789342.00 | T | C | NON_SYNONYMOUS_CODING | Q78R | MODERATE | 8 | 1 | 11.11 | 4 | 4 | 50 | 0.11086 |

### **Convergent molecular evolution in acute myeloid leukaemia**

Supplementary material

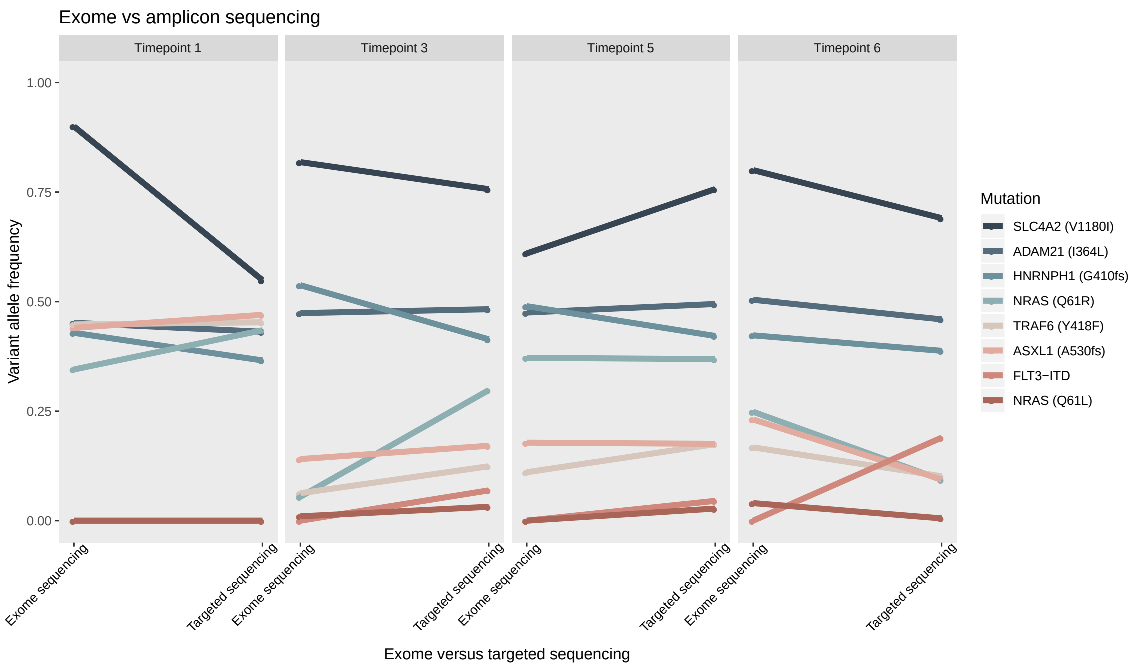


*Supplementary figure 4: Comparison of VAF from exome sequencing and the capture based targeted sequencing. The FLT3-ITD was not called in the exome sequencing experiment accounting for the variation of this variant.*

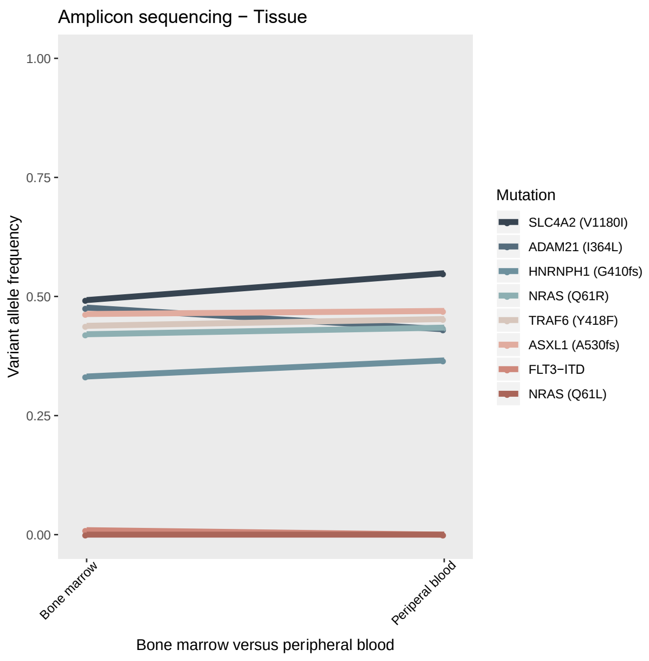


*Supplementary figure 5: Comparison of VAF between a bone marrow sample and a peripheral blood sample from TP1 as assessed by the targeted sequencing panel, demonstrating high concordance.*

#### Supplementary table 2: Targeted sequencing results

| Sample ID | *SLC4A2* (V1180I) | *ADAM21* (I364L) | *HNRNPH1* (G410fs) | *NRAS* (Q61R) | *TRAF6* (Y418F) | *ASXL1* (A530fs) | *FLT3*-ITD | *NRAS* (Q61L) |
|---|---|---|---|---|---|---|---|---|
| Control sample | 0.11324 | 0.10883 | 0.05096 | 0 | 0 | 0 | 0 | 0 |
| Timepoint 1 | 0.54893 | 0.43111 | 0.36585 | 0.4345 | 0.45278 | 0.46977 | 0 | 0 |
| Timepoint 2 | 0.16899 | 0.1495 | 0.18868 | 0.10898 | 0 | 0.07696 | 0 | 0 |
| Timepoint 3 | 0.7567 | 0.48261 | 0.41379 | 0.29794 | 0.12409 | 0.17104 | 0.06904 | 0 |
| Timepoint 4 | 0.849 | 0.48006 | 0.41935 | 0.08261 | 0 | 0.07518 | 0.15028 | 0 |
| Timepoint 5 | 0.75686 | 0.49437 | 0.42157 | 0.36863 | 0.17474 | 0.17546 | 0 | 0 |
| Timepoint 6 | 0.69056 | 0.45956 | 0.38764 | 0.09326 | 0.10147 | 0.09393 | 0.18936 | 0 |
| Timepoint 5 | 0.75686 | 0.49437 | 0.42157 | 0.36863 | 0.17474 | 0.17546 | 0 | 0 |
| CD11c- | 0.8017 | 0.49846 | 0.42045 | 0.15129 | 0.11801 | 0.09941 | 0.05036 | 0.13716 |
| CD11c+ | 0.71515 | 0.48557 | 0.3663 | 0.4168 | 0.15013 | 0.19136 | 0 | 0.06409 |
| CD19-CD7+ | 0.7788 | 0.47981 | 0.38344 | 0.25223 | 0.15963 | 0.185 | 0 | 0 |
| CD19+CD7- | 0.96901 | 0.47209 | 0.25 | 0 | 0 | 0 | 0.28989 | 0 |
| CD19+CD7+ | 0.99676 | 0.50063 | 0.4 | 0 | 0 | 0 | 0.27027 | 0 |
| CD3+ | 0.43283 | 0.32313 | 0.25828 | 0.21395 | 0.08086 | 0.06172 | 0 | 0 |
| Timepoint 1 - PDX a (5166 I) | 0 | 0.50976 | 0.54545 | 0.44843 | 0.45887 | 0.48173 | 0 | 0 |
| Timepoint 1 - PDX b (4745 I) | 0.919 | 0.53982 | 0.53846 | 0 | 0 | 0 | 0 | 0 |
| Timepoint 1 - PDX c (248 I) | 0.85185 | 0.6 | 0 | 0.37143 | 0 | 0 | 0 | 0 |
| Timepoint 1 - PDX d (4744 I) | 0.9625 | 0.26316 | 0 | 0 | 0 | 0 | 0 | 0.44928 |
| Timepoint 1 - PDX e (245 I) | NA | NA | NA | NA | NA | NA | NA | NA |
| Timepoint 1 - PDX f (247 I) | NA | NA | NA | NA | NA | NA | NA | NA |
| Timepoint 1 - PDX g (246 I) | NA | NA | NA | NA | NA | NA | NA | NA |
| Timepoint 1 - PDX h (249 I) | NA | NA | NA | NA | NA | NA | NA | NA |
| Timepoint 3 - PDX a (249 II) | 0.56911 | 0.2037 | 0 | 0.39785 | 0 | 0.0904 | 0 | 0.34409 |
| Timepoint 3 - PDX b (4744 II) | 0.72217 | 0.50333 | 0.40642 | 0.12877 | 0.10036 | 0.11719 | 0 | 0.37377 |
| Timepoint 3 - PDX c (246 II) | 0.64164 | 0.49695 | 0.51037 | 0.1409 | 0.14688 | 0.15472 | 0 | 0.36535 |
| Timepoint 3 - PDX d (245 II) | 0.71022 | 0.5 | 0.53631 | 0.15859 | 0.15008 | 0.16909 | 0 | 0.3507 |
| Timepoint 3 - PDX e (248 II) | 0.99625 | 0.51307 | 0.49558 | 0 | 0 | 0 | 0 | 0.54159 |
| Timepoint 3 - PDX f (246 NM) | 0 | 0 | 0 | 0 | 0 | 0 | 0 | 0 |
| Timepoint 3 - PDX g (4734 II) | NA | NA | NA | NA | NA | NA | NA | NA |
| Timepoint 3 - PDX h (4745 II) | NA | NA | NA | NA | NA | NA | NA | NA |
| Timepoint 5 - PDX a (5166 III) | 0.53366 | 0.48764 | 0.42581 | 0.42665 | 0.05351 | 0 | 0 | 0.12219 |
| Timepoint 5 - PDX b (246 III) | 0.57466 | 0.52176 | 0.49049 | 0.40288 | 0 | 0 | 0 | 0.1317 |
| Timepoint 5 - PDX c (245 III) | 0.47517 | 0.50595 | 0.45506 | 0.4508 | 0 | 0 | 0 | 0.06729 |
| Timepoint 5 - PDX d (245 NM) | 0.48922 | 0.49367 | 0.34211 | 0.48912 | 0 | 0 | 0 | 0 |
| Timepoint 5 - PDX e (247 II) | 0.73 | 0.50211 | 0.39 | 0.19872 | 0 | 0 | 0 | 0.33123 |
| Timepoint 5 - PDX f (249 NM) | 0.732 | 0.47298 | 0.47727 | 0.1939 | 0 | 0 | 0 | 0.31943 |
| Timepoint 5 - PDX g (4745 III) | 0.99813 | 0.4951 | 0.46341 | 0 | 0 | 0 | 0 | 0.52124 |
| Timepoint 5 - PDX h (4744 III) | 0.99384 | 0.49846 | 0.36681 | 0 | 0 | 0 | 0 | 0.51485 |
| Timepoint 6 - PDX a (4744 IIII) | 0.9965 | 0.4578 | 0.41155 | 0 | 0 | 0 | 0 | 0.5359 |
| Timepoint 6 - PDX b (4745 IIII) | 0.97086 | 0.49927 | 0.44019 | 0 | 0 | 0 | 0 | 0.51126 |
| Timepoint 6 - PDX c (248 NM) | 0.82625 | 0.5034 | 0.40945 | 0.14695 | 0.0611 | 0 | 0 | 0.36147 |
| Timepoint 6 - PDX d (246 IIII) | 0.53815 | 0.49315 | 0.44068 | 0.42811 | 0.09387 | 0.07878 | 0 | 0.09915 |
| Timepoint 6 - PDX e (245 IIII) | 0.67818 | 0.47895 | 0.44059 | 0.27601 | 0 | 0 | 0 | 0.23445 |
| Timepoint 6 - PDX f (4734 IIII) | 0.76108 | 0.49413 | 0.39147 | 0.1784 | 0 | 0 | 0 | 0.34431 |
| Timepoint 6 - PDX g (247 NM) | NA | NA | NA | NA | NA | NA | NA | NA |

Supplementary table 3:
Mass cytometry panel

| Target (Clone) | Mass | Phenograph/ UMAP |
|---|---|---|
| CD235a (HIR2) | 141 | - |
| CD19 (HIB19) | 142 | X |
| CD117 (104D2) | 143 | X |
| CD11b (ICRF44) | 144 | X |
| CD4 (RPAT4) | 145 | X |
| CD64 (10.1) | 146 | X |
| CD7 (CD7-6B7) | 147 | X |
| CD34 (581) | 148 | X |
| CD61 (VI-PL2) | 150 | - |
| CD123 (6H6) | 151 | X |
| CD13 (WM15) | 152 | X |
| CD62L (DREG-56) | 153 | X |
| CD45 (HI30) | 154 | - |
| CD183 (G025H7) | 156 | - |
| CD33 (WM53) | 158 | X |
| CD11c (Bu15) | 159 | X |
| CD14 (M5E2) | 160 | X |
| CD15 (W6D3) | 164 | - |
| CD16 (3G8) | 165 | - |
| CD24 (ML5) | 166 | - |
| CD38 (HIT2) | 167 | X |
| CD25 (2A3) | 169 | - |
| CD3 (UCHT1) | 170 | X |
| CD185 (51505)* | 171 | |
| HLA-DR (L243) | 174 | X |
| CD184 (12G5) | 175 | X |
| CD56 (CMSSB) | 176 | X |
| Iridium | 191 | |

The table presents the antibody panel used for mass cytometry staining and analysis. Clone of the antibody along with designation of type and mass number are listed for each antibody. The "Phenograhp/UMAP" column indicates whether the channel was included in the phenograph and UMAP analysis.

Supplementary table 4: Distribution of meta clusters across the four primary samples

| Sample | MC_01 | MC_02 | MC_03 | MC_04 | MC_05 | MC_06 | MC_07 | MC_08 | MC_09 | MC_10 | MC_11 | MC_12 |
|---|---|---|---|---|---|---|---|---|---|---|---|---|
| Timepoint 1 | 0.0 | 71.4 | 16.6 | 4.2 | 3.4 | 1.2 | 0.5 | 0.2 | 0.0 | 0.0 | 1.6 | 0.9 |
| Timepoint 3 | 14.9 | 0.0 | 51.6 | 1.5 | 1.5 | 2.0 | 11.3 | 0.1 | 11.2 | 0.0 | 5.9 | 0.0 |
| Timepoint 5 | 30.1 | 0.0 | 19.9 | 5.1 | 0.5 | 1.4 | 6.6 | 0.1 | 12.5 | 22.2 | 1.1 | 0.4 |
| Timepoint 6 | 59.3 | 0.0 | 23.3 | 1.4 | 0.3 | 1.2 | 5.1 | 0.0 | 9.0 | 0.0 | 0.0 | 0.4 |

The raw dual counts were arcsinh transformed with a co-factor of 5 and each of the four samples were clustered by PhenoGraph (K = 30) grouping individual cells based on pattern similarities and differences with regards to 18 surface markers. The marker selection was based on heterogeneity in signal across the samples for optimal separation. The analysis resulted in the delineation of 65 clusters, with an average of 16.25 clusters identified per sample (TP1: 20, TP3:13, TP5:18, TP6:14). To compare phenotypic composition across the various samples we preceded with a meta clustering approach (K = 3). This resulted in the identification of a total of 12 meta clusters (MC), outlining the range of diverse phenotypes derived from the index patient´s primary leukaemia samples. 6/12 MC were shared across all samples while 2/12 MC were identified only in a single sample (TP1:MC02, TP5:MC10).

#### Supplementary table 5: Overview of the 32 PDXes

| PDX ID | Cage | Cage-ID | Strain | Gender | Treat | Cells | OS (days) | Engraft. | CD45 | CD45 conf. | PCR | PCR conf. | TS | TS conf. | DSRT | Mass-cyt. | WES | CNA |
|---|---|---|---|---|---|---|---|---|---|---|---|---|---|---|---|---|---|---|
| TP1 - PDX a | 5166 | I | NSGS | Male | 15mg/kg | 1,7 xE06 | 285 | Yes | No | NA | S | OK | S | Yes | No | S & BM | S | No |
| TP1 - PDX b | 4745 | I | NSG | Female | 25mg/kg | 1,7 xE06 | 371 | Yes | S | OK | BM | No | BM | Yes | No | BM | No | No |
| TP1 - PDX c | 248 | I | NSGS | Male | 15mg/kg | 1,4 xE06 | 238 | Yes | NA | NA | S | No | S | Yes | No | No | No | No |
| TP1 - PDX d | 4744 | I | NSG | Female | 0 | 1,7 xE06 | 455 | Yes | NA | NA | BM | No | BM | Yes | No | No | No | No |
| TP1 - PDX e | 245 | I | NSGS | Female | 25mg/kg | 1,4 xE06 | 238 | No | S | No | BM | No | BM | No | No | No | No | No |
| TP1 - PDX f | 247 | I | NSGS | Male | 15mg/kg | 1,4 xE06 | 151 | No | BM | No | BM | No | BM | No | No | No | No | No |
| TP1 - PDX g | 246 | I | NSGS | Female | 25mg/kg | 1,4 xE06 | 342 | No | S | No | BM | No | No | NA | No | No | No | No |
| TP1 - PDX h | 249 | I | NSGS | Male | 15mg/kg | 1,4 xE06 | 174 | No | BM | No | BM | No | No | NA | No | No | No | No |
| TP3 - PDX a | 249 | II | NSGS | Male | 15mg/kg | 1,4 xE06 | 174 | Yes | S | OK | S | No | S | Yes | No | No | No | No |
| TP3 - PDX b | 4744 | II | NSG | Female | 0 | 1,7 xE06 | 455 | Yes | No | NA | S | OK | S | Yes | S | No | No | No |
| TP3 - PDX c | 246 | II | NSGS | Female | 25mg/kg | 1,4 xE06 | 222 | Yes | S | OK | S | OK | S | Yes | S | No | No | No |
| TP3 - PDX d | 245 | II | NSGS | Female | 25mg/kg | 1,4 xE06 | 214 | Yes | S | OK | S | OK | S | Yes | S | No | No | No |
| TP3 - PDX e | 248 | II | NSGS | Male | 15mg/kg | 1,4 xE06 | 230 | Yes | S | OK | S | OK | S | Yes | S | No | No | No |
| TP3 - PDX f | 246 | NM | NSGS | Female | 25mg/kg | 1,4 xE06 | 330 | Yes | S | OK | S | No | S | Yes | S | No | No | No |
| TP3 - PDX g | 4734 | II | NSGS | Male | 15mg/kg | 1,7 xE06 | 108 | Yes | S | OK | BM | OK | No | NA | No | S | No | No |
| TP3 - PDX h | 4745 | II | NSG | Female | 25mg/kg | 1,7 xE06 | 225 | Yes | No | NA | BM | OK | No | NA | No | S | No | No |
| TP5 - PDX a | 5166 | III | NSGS | Male | 15mg/kg | 2.2 xE06 | 108 | Yes | No | NA | S | OK | S | Yes | No | S & BM | No | S |
| TP5 - PDX b | 246 | III | NSGS | Female | 25mg/kg | 1,4 xE06 | 111 | Yes | S | OK | S | OK | S | Yes | S | No | No | No |
| TP5 - PDX c | 245 | III | NSGS | Female | 25mg/kg | 1,4 xE06 | 106 | Yes | S | OK | S | OK | S | Yes | S | No | No | No |
| TP5 - PDX d | 245 | NM | NSGS | Female | 25mg/kg | 1,4 xE06 | 105 | Yes | S | OK | S | OK | S | Yes | No | No | No | No |
| TP5 - PDX e | 247 | II | NSGS | Male | 15mg/kg | 1,4 xE06 | 151 | Yes | S | OK | S | OK | S | Yes | S | No | No | No |
| TP5 - PDX f | 249 | NM | NSGS | Male | 15mg/kg | 1,4 xE06 | 158 | Yes | S | OK | S | OK | S | Yes | No | No | No | No |
| TP5 - PDX g | 4745 | III | NSG | Female | 25mg/kg | 2.2 xE06 | 178 | Yes | S | OK | S | OK | S | Yes | S | S & BM | No | No |
| TP5 - PDX h | 4744 | III | NSG | Female | 0 | 2.2 xE06 | 234 | Yes | S | OK | S | OK | S | Yes | No | S | No | S |
| TP6 - PDX a | 4744 | IIII | NSG | Female | 0 | 3.2 xE06 | 166 | Yes | S | OK | S | OK | S | Yes | S | S & BM | No | No |
| TP6 - PDX b | 4745 | IIII | NSG | Female | 25mg/kg | 3.2 xE06 | 158 | Yes | S | OK | S | OK | S | Yes | S | S & BM | No | No |
| TP6 - PDX c | 248 | NM | NSGS | Male | 15mg/kg | 1,4 xE06 | 111 | Yes | S | OK | S | OK | S | Yes | No | No | No | No |
| TP6 - PDX d | 246 | IIII | NSGS | Female | 25mg/kg | 1,4 xE06 | 123 | Yes | S | OK | S | OK | S | Yes | S | No | No | No |
| TP6 - PDX e | 245 | IIII | NSGS | Female | 25mg/kg | 1,4 xE06 | 105 | Yes | S | OK | S | OK | S | Yes | S | No | No | No |
| TP6 - PDX f | 4734 | IIII | NSGS | Male | 15mg/kg | 3.2 xE06 | 108 | Yes | S | OK | S | OK | S | Yes | S | S & BM | S | No |
| TP6 - PDX g* | 247 | NM | NSGS | Male | 15mg/kg | 1,4 xE06 | 148 | NA | NA | NA | NA | NA | NA | NA | NA | NA | NA | No |

Treat: dosage of busulfan prior to injection of primary cells. OS: overall survival. Engraft: engraftment rate. S: Spleen. BM: Bone marrow. Conf: confirmation. TS: targeted sequencing. DSRT: drug sensitivity and resistance testing. Mass-cyt: mass-cytometry. WES: whole exome sequencing. CNA: copy number analysis.
*Mouse found dead in cage. Excluded from all analyses.

**Convergent molecular evolution in acute myeloid leukaemia**
Supplementary material

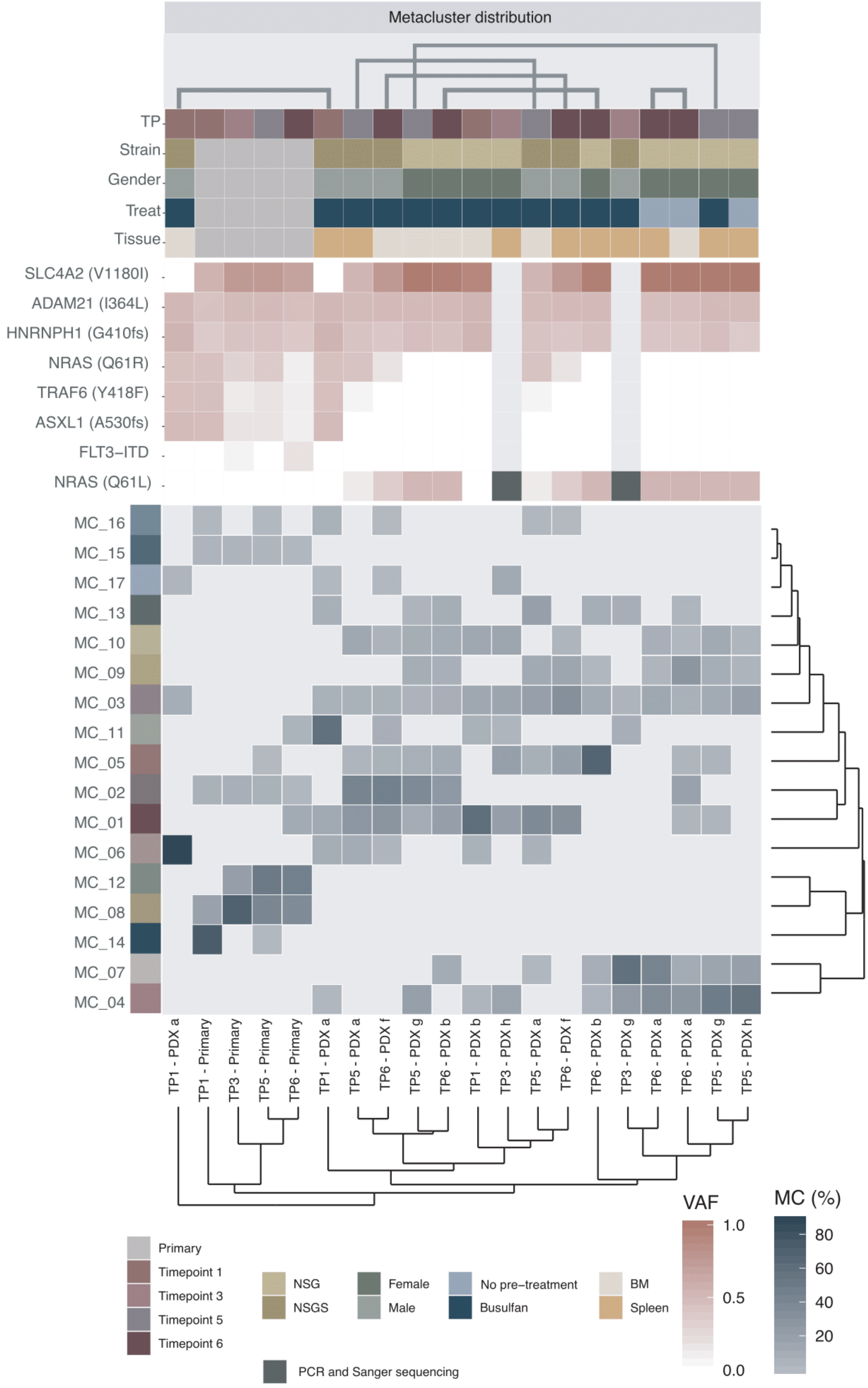


*Supplementary Figure 6: Hierarchical clustering (Euclidean distance, complete linkage) of the relative meta-cluster distribution. Blue colour indicates higher relative distribution and grey lower relative distribution. Empty tiles indicate that no cells were counted within that MC in the respective sample. The numerical values are presented in supplementary table 6. The MC names and colour indicators refer to the data presented in Figure **5A** and **5B**. The colour indicators at the top of the plot show sample characteristics, as presented in figure 3, indicating sample timepoint, strain, sex and pre-treatment. The red-white heat indicates VAF from the targeted sequencing experiment.*

#### Supplementary table 6: Distribution of meta clusters across the primary and PDX samples

| Sample | MC_01 | MC_02 | MC_03 | MC_04 | MC_05 | MC_06 | MC_07 | MC_08 | MC_09 | MC_10 | MC_11 | MC_12 | MC_13 | MC_14 | MC_15 | MC_16 | MC_17 |
|---|---|---|---|---|---|---|---|---|---|---|---|---|---|---|---|---|---|
| Timepoint 1 - Patient | 0.0 | 5.6 | 0.0 | 0.0 | 0.0 | 0.0 | 0.0 | 17.1 | 0.0 | 0.0 | 0.0 | 0.0 | 0.0 | 72.4 | 4.1 | 0.9 | 0.0 |
| Timepoint 1 - PDX a (5166 I) BM | 0.0 | 0.0 | 8.7 | 0.0 | 0.0 | 88.1 | 0.0 | 0.0 | 0.0 | 0.0 | 0.0 | 0.0 | 0.0 | 0.0 | 0.0 | 0.0 | 3.3 |
| Timepoint 1 - PDX a (5166 I) Spleen | 11.3 | 0.0 | 6.0 | 1.3 | 0.0 | 9.4 | 0.0 | 0.0 | 0.0 | 0.0 | 57.9 | 0.0 | 7.2 | 0.0 | 0.0 | 6.5 | 0.5 |
| Timepoint 1 - PDX b (4745 I) BM | 59.8 | 0.0 | 12.6 | 2.8 | 0.0 | 5.5 | 0.0 | 0.0 | 0.0 | 12.8 | 6.5 | 0.0 | 0.0 | 0.0 | 0.0 | 0.0 | 0.0 |
| Timepoint 3 - Patient | 0.0 | 7.4 | 0.0 | 0.0 | 0.0 | 0.0 | 0.0 | 68.9 | 0.0 | 0.0 | 0.0 | 20.2 | 0.0 | 0.0 | 3.5 | 0.0 | 0.0 |
| Timepoint 3 - PDX g (4734 II) Spleen | 0.0 | 0.0 | 3.5 | 23.5 | 0.0 | 0.0 | 57.0 | 0.0 | 0.0 | 0.0 | 7.9 | 0.0 | 8.1 | 0.0 | 0.0 | 0.0 | 0.0 |
| Timepoint 3 - PDX h (4745 II) Spleen | 20.0 | 0.0 | 15.4 | 15.7 | 19.4 | 0.0 | 0.0 | 0.0 | 0.0 | 15.0 | 3.3 | 0.0 | 0.0 | 0.0 | 0.0 | 0.0 | 11.1 |
| Timepoint 5 - Patient | 0.0 | 6.4 | 0.0 | 0.0 | 0.2 | 0.0 | 0.0 | 40.0 | 0.0 | 0.0 | 0.0 | 50.1 | 0.0 | 0.6 | 2.2 | 0.5 | 0.0 |
| Timepoint 5 - PDX a (5166 III) BM | 37.2 | 0.0 | 18.7 | 0.0 | 7.2 | 5.2 | 0.8 | 0.0 | 11.3 | 0.0 | 0.0 | 0.0 | 18.9 | 0.0 | 0.0 | 0.8 | 0.0 |
| Timepoint 5 - PDX a (5166 III) Spleen | 26.3 | 42.2 | 5.8 | 0.0 | 3.2 | 10.0 | 0.0 | 0.0 | 0.0 | 12.5 | 0.0 | 0.0 | 0.0 | 0.0 | 0.0 | 0.0 | 0.0 |
| Timepoint 5 - PDX g (4745 III) BM | 9.7 | 36.6 | 4.2 | 19.6 | 7.1 | 0.0 | 0.0 | 0.0 | 9.7 | 9.4 | 0.0 | 0.0 | 3.8 | 0.0 | 0.0 | 0.0 | 0.0 |
| Timepoint 5 - PDX g (4745 III) Spleen | 2.1 | 0.0 | 10.4 | 48.1 | 3.5 | 0.0 | 15.3 | 0.0 | 8.7 | 11.9 | 0.0 | 0.0 | 0.0 | 0.0 | 0.0 | 0.0 | 0.0 |
| Timepoint 5 - PDX h (4744 III) Spleen | 0.0 | 0.0 | 18.7 | 55.5 | 0.0 | 0.0 | 18.9 | 0.0 | 3.7 | 3.2 | 0.0 | 0.0 | 0.0 | 0.0 | 0.0 | 0.0 | 0.0 |
| Timepoint 6 - Patient | 11.0 | 1.2 | 0.0 | 0.0 | 0.0 | 0.0 | 0.0 | 36.1 | 0.0 | 0.0 | 4.6 | 45.7 | 0.0 | 0.0 | 1.5 | 0.0 | 0.0 |
| Timepoint 6 - PDX a (4744 IIII) BM | 2.4 | 18.2 | 5.9 | 27.7 | 2.7 | 0.0 | 10.9 | 0.0 | 27.4 | 1.5 | 0.0 | 0.0 | 3.3 | 0.0 | 0.0 | 0.0 | 0.0 |
| Timepoint 6 - PDX a (4744 IIII) Spleen | 0.0 | 0.0 | 13.5 | 35.1 | 0.0 | 0.0 | 42.0 | 0.0 | 2.6 | 6.8 | 0.0 | 0.0 | 0.0 | 0.0 | 0.0 | 0.0 | 0.0 |
| Timepoint 6 - PDX b (4745 IIII) BM | 19.6 | 24.6 | 7.4 | 0.0 | 10.2 | 0.0 | 11.0 | 0.0 | 6.9 | 10.3 | 0.0 | 0.0 | 10.0 | 0.0 | 0.0 | 0.0 | 0.0 |
| Timepoint 6 - PDX b (4745 IIII) Spleen | 0.0 | 0.0 | 10.5 | 3.9 | 66.5 | 0.0 | 8.6 | 0.0 | 1.7 | 0.0 | 0.0 | 0.0 | 8.8 | 0.0 | 0.0 | 0.0 | 0.0 |
| Timepoint 6 - PDX f (4734 IIII) BM | 28.0 | 45.7 | 6.4 | 0.0 | 5.2 | 2.5 | 0.0 | 0.0 | 0.0 | 4.2 | 7.2 | 0.0 | 0.0 | 0.0 | 0.0 | 0.3 | 0.4 |
| Timepoint 6 - PDX f (4734 IIII) Spleen | 31.9 | 0.0 | 31.3 | 0.0 | 19.0 | 0.0 | 0.0 | 0.0 | 14.9 | 2.6 | 0.0 | 0.0 | 0.0 | 0.0 | 0.0 | 0.3 | 0.0 |

All of the data, including the events collected from the four primary samples, were then reanalysed following the same analytic approach applied for the assessment of the primary samples. Due to substantial variation in acquired events across the 20 files subsampling was performed with a maximum event count of 10 000 (range: 2 221-10 000) per file. Individual PhenoGraph clustering (K: 30) resulted in identification of 228 clusters with an average of 11.4 clusters identified per sample (range: 8-16). Meta clustering (K:5) resulted in identification of 17 MC, with each sample characterised by a relative contribution of 3-9 MC (total:128, mean per sample 6.4) at a relative contribution ranging from 0.16 to 88.08% (mean: 15.6%).

#### Supplementary table 7: Expression of human CD45

| PDX ID | CD45 | CD45 confirm | Human | Mouse | Fraction |
|---|---|---|---|---|---|
| Timepoint 1 - PDX a (5166 I) | No | NA | NA | NA | NA |
| Timepoint 1 - PDX b (4745 I) | Spleen | OK | 348 | 26536 | 1.3 |
| Timepoint 1 - PDX c (248 I) | No | NA | NA | NA | NA |
| Timepoint 1 - PDX d (4744 I) | No | NA | NA | NA | NA |
| Timepoint 1 - PDX e (245 I) | Spleen | No | 2 | 47854 | 0.0 |
| Timepoint 1 - PDX f (247 I) | Bone marrow | No | 3 | 18706 | 0.0 |
| Timepoint 1 - PDX g (246 I) | Spleen | No | 38 | 55297 | 0.1 |
| Timepoint 1 - PDX h (249 I) | Bone marrow | No | 46 | 185 | 19.9 |
| Timepoint 3 - PDX a (249 II) | Spleen | OK | 559 | 20515 | 2.7 |
| Timepoint 3 - PDX b (4744 II) | No | NA | NA | NA | NA |
| Timepoint 3 - PDX c (246 II) | Spleen | OK | 20604 | 366 | 98.3 |
| Timepoint 3 - PDX d (245 II) | Spleen | OK | 21232 | 1671 | 92.7 |
| Timepoint 3 - PDX e (248 II) | Spleen | OK | 19060 | 1098 | 94.6 |
| Timepoint 3 - PDX f (246 NM) | Spleen | OK | 14650 | 4155 | 77.9 |
| Timepoint 3 - PDX g (4734 II) | Spleen | OK | 1468 | 22665 | 6.1 |
| Timepoint 3 - PDX h (4745 II) | No | NA | NA | NA | NA |
| Timepoint 5 - PDX a (5166 III) | No | NA | NA | NA | NA |
| Timepoint 5 - PDX b (246 III) | Spleen | OK | 16240 | 2170 | 88.2 |
| Timepoint 5 - PDX c (245 III) | Spleen | OK | 3998 | 1012 | 79.8 |
| Timepoint 5 - PDX d (245 NM) | Spleen | OK | 4593 | 5249 | 46.7 |
| Timepoint 5 - PDX e (247 II) | Spleen | OK | 18991 | 1321 | 93.5 |
| Timepoint 5 - PDX f (249 NM) | Spleen | OK | 714 | 1921 | 27.1 |
| Timepoint 5 - PDX g (4745 III) | Spleen | OK | 20588 | 475 | 97.7 |
| Timepoint 5 - PDX h (4744 III) | Spleen | OK | 22744 | 247 | 98.9 |
| Timepoint 6 - PDX a (4744 IIII) | Spleen | OK | 20844 | 229 | 98.9 |
| Timepoint 6 - PDX b (4745 IIII) | Spleen | OK | 328 | 40 | 89.1 |
| Timepoint 6 - PDX c (248 NM) | Spleen | OK | 7292 | 12572 | 36.7 |
| Timepoint 6 - PDX d (246 IIII) | Spleen | OK | 15997 | 2220 | 87.8 |
| Timepoint 6 - PDX e (245 IIII) | Spleen | OK | 1576 | 537 | 74.6 |
| Timepoint 6 - PDX f (4734 IIII) | Spleen | OK | 17508 | 1988 | 89.8 |
| Timepoint 6 - PDX g (247 NM) | NA | NA | NA | NA | NA |
| Timepoint 6 - PDX h (NA NA) | NA | NA | NA | NA | NA |

**Convergent molecular evolution in acute myeloid leukaemia**
Supplementary material

#### Supplementary table 8: Overview of drugs in the drug sensitivity and resistance screen

| Drug Class | Drug name | Drug subclass | FIMM ID |
|---|---|---|---|
| A. Conv. Chemo | Bortezomib | Proteasome inhibitor (26S subunit) | FIMM000287-002 |
| A. Conv. Chemo | Carfilzomib | Proteasome inhibitor (20S subunit) | FIMM100346-001 |
| A. Conv. Chemo | Chloroquine | Antimalaria agent; chemo/radio sensitizer | FIMM100371-002 |
| A. Conv. Chemo | Clofarabine | Antimetabolite; Purine analog | FIMM000321-002 |
| A. Conv. Chemo | Cytarabine | Antimetabolite, interferes with DNA synthesis | FIMM000406-003 |
| A. Conv. Chemo | Etoposide | Topoisomerase II inhibitor | FIMM023814-002 |
| A. Conv. Chemo | Fludarabine | Antimetabolite; Purine analog | FIMM023828-002 |
| A. Conv. Chemo | Gemcitabine | Antimetabolite; Nucleoside analog | FIMM023798-002 |
| A. Conv. Chemo | Hydroxyurea | Antineoplastic agent | FIMM000983-003 |
| A. Conv. Chemo | Idarubicin | Topoisomerase II inhibitor | FIMM000173-003 |
| A. Conv. Chemo | Mitoxantrone | Topoisomerase II inhibitor | FIMM000490-004 |
| A. Conv. Chemo | Omacetaxine | Protein synthesis inhib (80 S ribosome) | FIMM100379-001 |
| A. Conv. Chemo | Paclitaxel | Mitotic inhibitor, taxane microtubule stabilizer | FIMM000491-003 |
| A. Conv. Chemo | Topotecan | Topoisomerase I inhibitor. Camptothecin analog | FIMM000556-005 |
| A. Conv. Chemo | Vincristine | Mitotic inhibitor. Vinca alkaloid microtubule depolymerizer | FIMM000337-003 |
| B. Kinase inhibitor | Afatinib | EGFR inhibitor | FIMM003710-001 |
| B. Kinase inhibitor | Alisertib | Aurora A inhibitor | FIMM003761-001 |
| B. Kinase inhibitor | Alvocidib | CDK inhibitor | FIMM003725-001 |
| B. Kinase inhibitor | Axitinib | VEGFR, PDGFR, KIT inhibitor | FIMM003778-001 |
| B. Kinase inhibitor | Bosutinib | Abl, Src inhibitor | FIMM003780-002 |
| B. Kinase inhibitor | Crenolanib | PDGFRA and PDGFRB inhibitor | FIMM109464-001 |
| B. Kinase inhibitor | Dasatinib | Abl, Src, Kit, EphR... Inhibitor | FIMM003775-001 |
| B. Kinase inhibitor | Dinaciclib | CDK inhibitor | FIMM109460-001 |
| B. Kinase inhibitor | Erlotinib | EGFR inhibitor | FIMM000183-003 |
| B. Kinase inhibitor | Midostaurin | Flt3, PKC, PKA, S6K and EGFR inhibitor | FIMM100362-001 |
| B. Kinase inhibitor | Momelotinib | JAK1 & 2 inhibitor | FIMM109441-001 |
| B. Kinase inhibitor | Nintedanib | FLT3, VEGFR, PDGFR, FGFR inhibitor | FIMM100389-001 |
| B. Kinase inhibitor | Ponatinib | FLT3, Broad TK inhibitor | FIMM003730-001 |
| B. Kinase inhibitor | Regorafenib | B-Raf, c-Kit, VEGFR2 inhibitor | FIMM100364-001 |
| B. Kinase inhibitor | Ruxolitinib | JAK1&2 inhibitor | FIMM003756-001 |
| B. Kinase inhibitor | Sunitinib | FLT3, Broad TK inhibitor | FIMM023820-002 |
| B. Kinase inhibitor | Trametinib | MEK1/2 inhibitor | FIMM003751-001 |
| B. Kinase inhibitor | Vemurafenib | B-Raf(V600E) inhibitor | FIMM003767-001 |
| B. Kinase inhibitor | Volasertib | PLK1 inhibitor | FIMM100375-001 |
| C. Rapalog | Everolimus | binds FKBP12, causes inhibition of mTORC1 | FIMM003755-001 |
| D. Immunomodulatory | Dexamethasone | Immunosuppresant; glucocorticoid | FIMM000227-002 |
| D. Immunomodulatory | Methylprednisolone | Immunosuppressant | FIMM000484-002 |
| E. Differentiating/ epigenetic modifier | Azacitidine | Nucleoside analog DNA methyl transferase inhibitor | FIMM023825-003 |
| E. Differentiating/ epigenetic modifier | Belinostat | HDAC inhibitor | FIMM003744-001 |
| E. Differentiating/ epigenetic modifier | Olaparib | PARP inhibitor | FIMM003790-001 |
| E. Differentiating/ epigenetic modifier | Panobinostat | HDAC inhibitor | FIMM003783-001 |
| E. Differentiating/ epigenetic modifier | Rucaparib | PARP inhibitor | FIMM003736-001 |
| E. Differentiating/ epigenetic modifier | Tipifarnib | Farnesyltransferase inhibitor | FIMM003728-001 |
| E. Differentiating/ epigenetic modifier | Tretinoin | Retinoic acid receptor agonist | FIMM000249-004 |
| E. Differentiating/ epigenetic modifier | Vorinostat | HDAC inhibitor | FIMM003787-001 |
| G. Apoptotic modulator | Navitoclax | Bcl-2/Bcl-xL inhibitor | FIMM003707-001 |
| H. Metabolic modifier | Methotrexate | Antimetabolite; Anti-folate agent | FIMM000649-004 |
| H. Metabolic modifier | Pravastatin | HMG CoA reductase inhibitor | FIMM003569-002 |
| J. NSAID | Celecoxib | Selective COX-2 inhibitor | FIMM000298-004 |
| X. Other | Anagrelide | PDE-3, PLA2 inhibitor | FIMM000171-004 |
| X. Other | Mepacrine | Unclear. PLA2 inhibitor. NF-kB inhibitor, p53 activator | FIMM000528-005 |
| X. Other | Metformin | Unknown | FIMM109466-001 |
| X. Other | Tosedostat | Aminopeptidase inhibitor | FIMM100402-001 |
| X. Other | Vismodegib | Smothened (Hh) inhibitor | FIMM003793-001 |

**Convergent molecular evolution in acute myeloid leukaemia**
Supplementary material

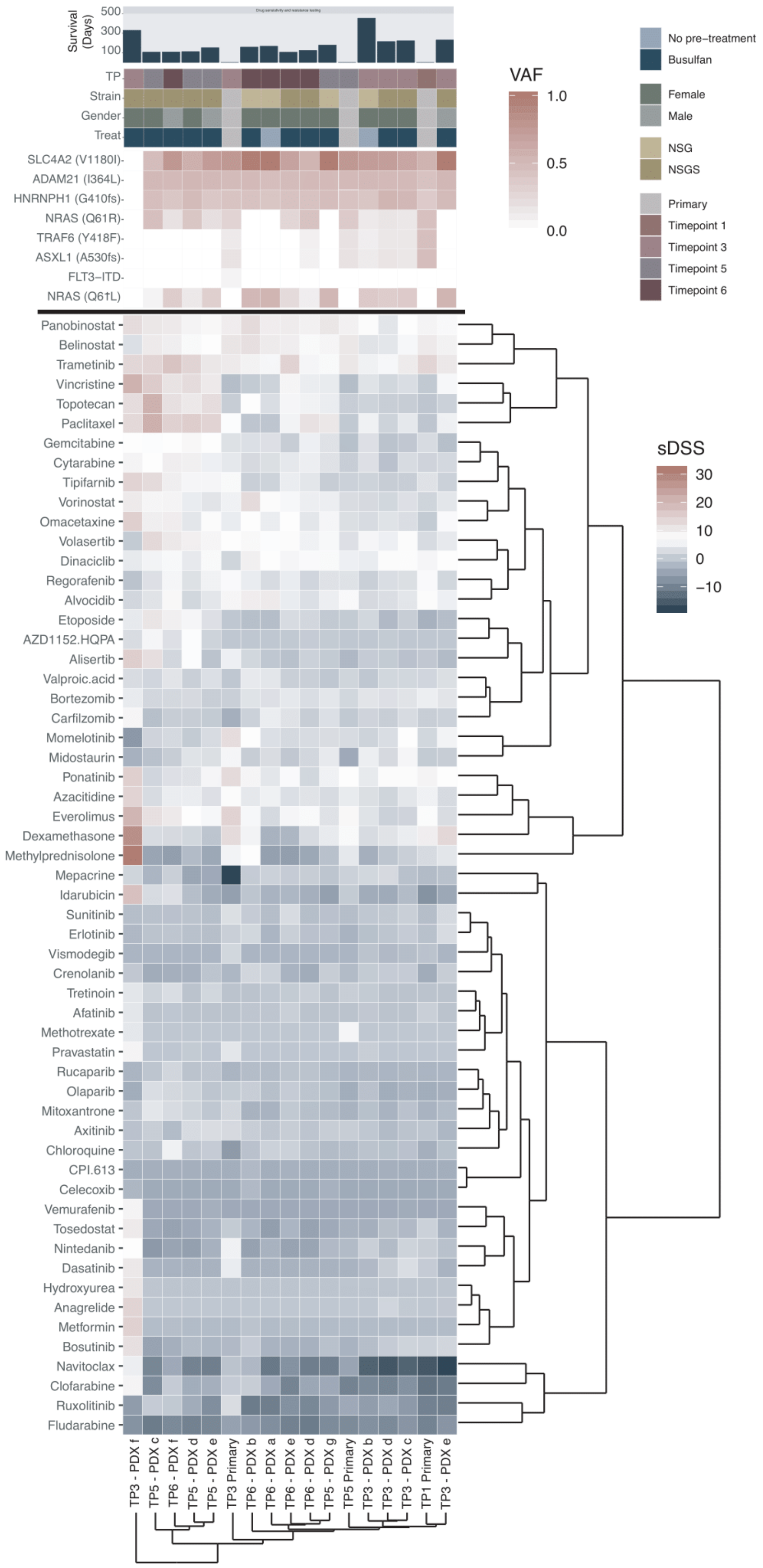


*Supplementary Figure 7: Hierarchical clustering (Euclidean distance, complete linkage) of drug sensitivity scores of 58 anti-cancer compounds in primary and PDX-derived samples. The colour gradient represents the sDSS values (DSS-median DSS of two normal cryopreserved and thawed BM samples). Red signifies a high sDSS and sensitivity, while blue signifies a negative sDSS and a relative resistance to the drug. The indicators above the heatmap show sample identity, as presented in figure 3. All PDX samples are derived from spleen.*

### Supplementary table 9: Overview of drugs scored as sensitive (sDSS ≥5) in at least half of all screened samples

| Drug name | Belinostat | Dinaciclib | Everolimus | Omacetaxine | Paclitaxel | Panobinostat | Ponatinib | Topotecan | Trametinib | Volasertib | Vorinostat |
|---|---|---|---|---|---|---|---|---|---|---|---|
| Drug subclass | HDAC inhibitor | CDK inhibitor | binds FKBP12, causes inhibition of mTORC1 | Protein synthesis inhib (80 S ribosome) | Mitotic inhibitor, taxane microtubule stabilizer | HDAC inhibitor | FLT3, Broad TK inhibitor | Topoisomerase I inhibitor. Camptothecin analog | MEK1/2 inhibitor | PLK1 inhibitor | HDAC inhibitor |
| Primary TP1 | 9,97 | 5,83 | 7,19 | 0,54 | -0,05 | 8,23 | 8,42 | -0,09 | 13,65 | 2,71 | 1,85 |
| Primary TP3 | 10,87 | 1,90 | 14,30 | 4,88 | 6,61 | 9,51 | 12,63 | 0,88 | 9,81 | 6,75 | 5,56 |
| Primary TP5 | 10,45 | 5,27 | 7,82 | 3,23 | 0,28 | 8,91 | 6,84 | 1,10 | 10,35 | 3,38 | 2,94 |
| Timepoint 3 - PDX d (245 II) spleen | 4,15 | 6,55 | 1,40 | 3,55 | 1,65 | 4,05 | 7,35 | 1 | 6,05 | 5,20 | 2,15 |
| Timepoint 3 - PDX f (246 NM) spleen | 3,35 | 4,65 | 19,90 | 14,95 | 12,95 | 12,35 | 14,85 | 12,20 | 13,35 | 0,50 | 9,85 |
| Timepoint 3 - PDX c (246 II) spleen | 6,15 | 7,15 | 1,80 | 3,15 | 4,35 | 6,75 | 7,25 | 0,80 | 9,75 | 7,30 | 3,45 |
| Timepoint 5 - PDX e (247 II) spleen | 6,95 | 7,45 | 6,10 | 7,35 | 14,05 | 7,85 | 5,85 | 12,20 | 10,25 | 8,10 | 3,95 |
| Timepoint 3 - PDX e (248 II)spleen | 8,65 | 6,85 | 2,90 | 6,45 | 5,35 | 6,35 | 7,05 | 1,50 | 10,15 | 4,60 | 4,25 |
| Timepoint 3 - PDX b (4744 II) spleen | 3,75 | 4,85 | 3,60 | 2,65 | 5,45 | 7,35 | 4,65 | 1,70 | 5,45 | 5,90 | 2,45 |
| Timepoint 5 - PDX d (245 NM) spleen | 9,15 | 5,75 | 11,00 | 7,55 | 18,35 | 11,05 | 1,25 | 14,50 | 15,75 | 9,50 | 7,55 |
| Timepoint 5 - PDX c (245 III) spleen | 9,85 | 5,75 | 11,60 | 9,15 | 21,15 | 10,35 | 1,95 | 20,80 | 13,75 | 13 | 8,05 |
| Timepoint 5 - PDX g (4745 III) spleen | 8,05 | 7,55 | 3,70 | 5,95 | 10,25 | 10,55 | 5,55 | 5 | 7,55 | 6,20 | 5,15 |
| Timepoint 6 - PDX e (245 IIII) spleen | 9,25 | 5,95 | 5,90 | 6,95 | 5,65 | 9,65 | 6,45 | 5,90 | 14,25 | 6,50 | 6,15 |
| Timepoint 6 - PDX d (246 IIII) spleen | 7,45 | 6,45 | 7,50 | 4,25 | 11,35 | 8,75 | 2,15 | 5,60 | 5,95 | 5,50 | 5,15 |
| Timepoint 6 - PDX f (4734 IIII) spleen | 9,15 | 6,25 | 10,10 | 9,35 | 14,55 | 9,95 | 3,45 | 11,60 | 17,15 | 10,10 | 8,35 |
| Timepoint 6 - PDX a (4744 IIII) spleen | 9,75 | 6,75 | 2,50 | 5,05 | -0,05 | 9,35 | 4,05 | 3,10 | 6,05 | 6,60 | 7,25 |
| Timepoint 6 - PDX b (4745 IIII) spleen | 11,75 | 8,95 | 6,10 | 6,95 | -0,05 | 12,75 | 8,15 | 7,10 | 8,65 | 7,60 | 12,55 |
